# The development and validation of a prognostic model to predict relapse in adults with remitted depression in primary care: secondary analysis of pooled individual participant data from multiple studies

**DOI:** 10.1101/2024.06.25.24309402

**Authors:** Andrew S Moriarty, Lewis W Paton, Kym IE Snell, Lucinda Archer, Richard D Riley, Joshua EJ Buckman, Carolyn A Chew-Graham, Simon Gilbody, Shehzad Ali, Stephen Pilling, Nick Meader, Bob Phillips, Peter A Coventry, Jaime Delgadillo, David A Richards, Chris Salisbury, Dean McMillan

## Abstract

**Background:** Relapse of depression is common and contributes to the overall associated morbidity and burden. We lack evidence-based tools to estimate an individual’s risk of relapse after treatment in primary care, which may help us more effectively target relapse prevention.

**Objective:** Develop and validate a prognostic model to predict risk of relapse of depression in primary care.

**Methods:** Multilevel logistic regression models were developed, using individual participant data from seven primary care-based studies (n=1244), to predict relapse of depression. The model was internally validated using bootstrapping and generalisability was explored using internal-external cross-validation.

**Findings:** Residual depressive symptoms [Odds ratio (OR): 1.13 (95% CI: 1.07-1.20), p<0.001] and baseline depression severity [OR: 1.07 (1.04-1.11), p<0.001] were associated with relapse. The validated model had low discrimination [C-statistic 0.60 (0.55-0.65)] and miscalibration concerns [calibration slope 0.81 (0.31-1.31)]. On secondary analysis, being in a relationship was associated with reduced risk of relapse [OR: 0.43 (0.28-0.67), p<0.001]; this remained statistically significant after correction for multiple significance testing.

**Conclusions:** We cannot currently predict risk of depression relapse with sufficient accuracy in a primary care setting, using routinely recorded measures. Relationship status warrants further research to explore its role as a prognostic factor for relapse.

**Clinical implications:** Until we can accurately stratify patients according to risk of relapse, a universal approach to relapse prevention may be most beneficial, either during acute phase treatment or post-remission. Where possible, this could be guided by the presence or absence of known prognostic factors (e.g. residual depressive symptoms) and targeted towards these.

**What is already known on this topic:** Relapse contributes to the morbidity and burden associated with depression and, while there is robust research confirming predictors of relapse, individualised risk prediction is a challenge.

**What this study adds:** We found that it is not possible to accurately predict individualised risk of relapse using prognostic factors that are routinely collected and available in primary care. We found evidence to suggest that relationship status (not being in a relationship) is associated with increased risk of relapse and warrants confirmatory prognostic factor research.

**How this study might affect research, practice or policy:** Future prognosis research in this area should focus on exploring the feasibility of routinely measuring and documenting additional prognostic factors in primary care (for example, adverse childhood events, relationship status and social support) and including these in prognostic models. Until we can more accurately identify individuals at increased risk of relapse, commonly used acute-phase treatments could be optimised to better prepare for and mitigate the risk of relapse and there is a need for brief, scalable relapse prevention interventions that could be provided more widely.

## 1. BACKGROUND

Depression is the leading cause of disability worldwide(1); the vast majority of adults seeking treatment for depression are managed in primary care(2). Relapse is common, with around half of people experiencing a relapse within one year of reaching remission(3). This high relapse rate contributes to the overall morbidity and burden associated with depression(4).

An ability to predict an individual patient’s risk of relapse after an episode of depression might assist clinicians in targeting relapse prevention interventions toward those at greatest risk. Well-established prognostic factors associated with increased risk of relapse are: residual depressive symptoms; previous depressive episodes; childhood maltreatment; comorbid anxiety; neuroticism; younger age of first onset, and rumination(5). While the presence or absence of these prognostic factors can help refine estimates of overall prognosis to particular sub-groups, they do not effectively aid risk-stratification at the individual level. Sub-grouping methods have been used to predict average risk of relapse for groups of people with different combinations (or profiles) of prognostic factors(6). However, individualised outcome prediction is best shaped using multiple prognostic factors in combination, in the form of multivariable prognostic models(7).

A systematic review of prognostic models, undertaken by our group, identified twelve studies of relapse prediction models(8). The majority were at high overall risk of bias (the most significant limitations being inadequate sample size, inappropriate handling of missing data, and calibration or discrimination not reported). The developed models either demonstrated insufficient predictive performance on reported validation by the study authors, or they could not be feasibly implemented in a primary care setting due to the large number and type of included predictors. We concluded that we currently lack evidence-based tools to assist clinicians with risk prediction of depressive relapse in any clinical setting, and that new models are required to give accurate risk predictions in primary care settings.

## 2. OBJECTIVE

To develop and validate a prognostic model, for use in clinical primary care settings, to predict risk of relapse in adults with remitted depression(9).

## 3. METHODS

The methods align with the PROGnosis RESearch Strategy (PROGRESS) recommendations(7) and the study is reported according to the Transparent Reporting of a multivariable prediction model for Individual Prognosis or Diagnosis (TRIPOD-Cluster) guidance(10). A Patient Advisory Group (PAG) contributed to this study, including selecting predictors, definition of outcome, target patient population and clinical application. The study was registered prospectively (ClinicalTrials.gov: NCT04666662). Further methodological details are available in our protocol paper(9).

### 3.1. Source of data, participants and setting

We formed the “PREDICTR” dataset from combined individual participant data (IPD) from UK primary care-based studies(9), identified through a literature search and a review of the NIHR trials registry. Authors were asked to share data if studies included adult patients (18 years and over) with depression, measured using the Patient Health Questionnaire (PHQ-9) at a minimum of three time-points (to identify depression, remission, relapse/no relapse). We excluded studies in patient groups with significant psychiatric comorbidity and feasibility studies. The PREDICTR dataset is derived from all arms (control and intervention) of six RCTs of primary care-based interventions for depression (CADET, CASPER Plus, COBRA, Healthlines Depression, REEACT, and REEACT-2) and one observational cohort study (WYLOW) (Supplemental Table 1.1, Figure 1.1).

### 3.2. Start-point (remission)

Participants were in remission at the point of prediction. Participants must have had case-level depression at baseline [PHQ-9 score of 10 or more(11)], and (at 4 months after trial baseline): i) a post-treatment PHQ-9 score below the established cut-off of 10 [consistent with clinical recovery(3,12)] and ii) an improvement of ≥5 points on the PHQ-9 since depression diagnosis [which aligns with an established reliable change index to identify those with “reliable improvement”(13)].

### 3.3. End-point/outcome (relapse)

We coded participants as relapsed if they fulfilled the following criteria within 6-8 months post-remission: i) PHQ-9 score above the diagnostic cut-off (10 or more) and ii) ≥5 points greater than their symptom score at the time of remission. This is also consistent with an established criteria for reliable and clinically significant deterioration(13).

### 3.4. Predictors

We identified predictors *a priori,* following a literature review and consensus within the multidisciplinary research team and the PAG. We included predictors that would currently be routinely available in primary care settings at the intended moment of prediction.

#### 3.4.1. Predictors in primary analysis

The following variables have robust evidence for their role as relapse predictors(5,14) and were included in the model:

- Residual depressive symptoms (PHQ-9 score at remission (0–9); continuous variable);
- Previous episodes of depression (dichotomous predictor (0=no previous episodes, 1=one or more previous episodes;
- Comorbid anxiety (measured using the GAD-7(15) in six of the seven studies and the Clinic Interview Schedule-Revised (CIS-R)(16) in REEACT (see Supplemental Table 1.2). These measures were combined to create a composite score (z-score), modelled as a continuous predictor;
- Baseline severity of depressive symptoms (continuous predictor; PHQ-9 score at baseline (pre-treatment));
- RCT Intervention: to control for the presence of interventions within the RCTs, we coded the presence or absence of an effective intervention (based on the results of the RCT) as a dichotomous variable. This predictor was intended to control for the intervention as part of the model building process only; when making predictions in real-world primary care this predictor would always be set to zero (i.e., no experimental intervention present).

#### 3.4.2. Exploratory predictors

These are less well-evidenced predictors of relapse and were included as part of an exploratory secondary analysis: age; gender; ethnicity; relationship status; multimorbidity (two or more long-term physical or mental health conditions, excluding depression and comorbid anxiety); employment status (unemployment being those of working age who do not have a job and are actively seeking one); and current antidepressant use(5,14,17).

### 3.5. Sample size

We used the *pmsampsize* package (18) to calculate the required minimum sample size of 722, with 145 events (see protocol for details(9)); our actual sample size (n=1244; 261 events) exceeded this.

### 3.6. Statistical analysis methods

#### 3.6.1. Data pre-processing

Anonymised data were transferred and stored securely and harmonised in line with the pre-specified harmonisation procedure (Supplemental Table 1.3).

#### 3.6.2. Data integrity checks (risk of bias)

Data were summarised and checked against publications for key features such as number of participants (total and in each study arm), demographics, primary outcomes of study, relapse rates and missing data. Validity of data values were checked on data inspection and irregularities clarified through communication with the original authors. Risk of bias assessment was undertaken using the participants, predictors and outcome domains of PROBAST(19).

#### 3.6.3. Missing data

Missing data were handled using multiple imputation with chained equations (MICE), under a missing at random assumption(20). Missing values were imputed based on the values of other predictors and the outcome, using linear models for continuous predictors (residual symptoms, severity, comorbid anxiety) and logistic models for binary predictors [number of previous episodes, RCT Intervention, outcome (relapse/no relapse)]. Imputation was undertaken for each study separately, preserving the clustering of participants within studies and any between-study heterogeneity in predictor effects and outcome prevalence. Each imputed dataset was then analysed separately using the same statistical methods, and the estimates were combined using Rubin’s rules, to produce an overall estimate and measure of uncertainty of each regression coefficient and model performance measures(10). We used thirty imputations, based on the maximum percentage of participants with one or more missing values across all individual studies(20).

#### 3.6.4. Model development (primary analysis)

Multilevel multivariable logistic regression models were built to model the relationship of the predictors with the binary outcome (relapse/no relapse), forcing in all predictors. Model parameters were estimated via unpenalized maximum likelihood estimation, and then penalised post-estimation using a uniform shrinkage factor. The modelling preserved the clustering of participants within studies, with a random effect on the intercept, a random intervention effect, and allowing for between-study correlation in these effects. We explored non-linear relationships in the continuous variables using multivariable fractional polynomials (MFPs)(7).

Predictive performance statistics (C-statistic for discrimination; calibration slope and calibration-in-the-large) were calculated for the final developed model, first within each cluster in turn and then pooled using random effects meta-analysis to summarise the model’s performance across clusters with estimates of the pooled average and 95% confidence intervals. Prediction intervals were constructed to estimate the model’s likely performance in new but similar settings(10). Calibration was also assessed visually by producing calibration plots with smooth calibration curves.

#### 3.6.5. Model validation

The optimism of the developed model was measured using non-parametric bootstrapping. One hundred bootstrap samples (each stratified by study) were produced from the original dataset. Within each bootstrap sample, the same modelling procedures were used as for model development. The model estimated using each bootstrap sample was then applied in both the same bootstrap sample (‘apparent performance’) and in the original (imputed) dataset (‘test performance’). Each time, average performance measures were calculated by pooling within-study statistics using meta-analysis, as above.

Optimism was calculated as the difference between apparent and test performance; this process was repeated one hundred times and the average difference between the bootstrap (apparent) and test performance for each performance statistic provided the estimate of overall optimism for that statistic. Optimism-adjusted performance statistics (C-statistic, calibration slope and calibration-in-the-large) were subsequently derived. The uniform shrinkage factor (in this study, the optimism-adjusted calibration slope) was applied to all of the original estimated beta coefficients (to shrink them toward zero to address overfitting) to produce a penalised logistic regression model. Finally, the intercept was re-estimated (whilst constraining the penalised predictor effects at their shrunken value) to ensure overall calibration was maintained. This formed the final model. Generalisability of the model and between-study heterogeneity in model performance was assessed using internal-external cross-validation (IECV)(21) (see Supplemental Materials 4 for procedure).

#### 3.6.6. Sensitivity analysis

To understand the impact of including a composite measure of comorbid anxiety calculated from both GAD-7 and CIS-R, a sensitivity analysis was performed measuring predictive performance statistics when omitting REEACT and using only GAD-7 as the measure of comorbid anxiety, rather than a z-score.

#### 3.6.7. Secondary (exploratory) analyses

Univariable analyses were performed to evaluate the unadjusted association between each predictor variable and the outcome variable. Where univariable analysis found statistically significant associations (after accounting for multiple significance testing), the model was refit using all of the original included predictors plus the additional exploratory predictor, to explore the impact on model predictive performance (using only studies in which the exploratory predictor was available).

### 3.7. Ethics approval

The University of York’s Health Sciences Research Governance Committee confirmed that this study was exempt from full ethical approval as it entailed the secondary analysis of anonymised data from studies that had already received ethical approval.

## 4. RESULTS

### 4.1. Summary of data

Table 1 presents the descriptive statistics for the IPD. Supplemental Table 2.1 summarises the missing data. Risk of bias and concerns around applicability were low (Supplemental Table 2.4).

**Table 1:** Descriptive statistics for IPD.

| Variable | Study |  |  |  |  |  |  |  |
| --- | --- | --- | --- | --- | --- | --- | --- | --- |
|  | CADET | CASPER Plus | COBRA | Healthlines Depression | REEACT | REEACT-2 | WYLOW | Combined PREDICTR IPD dataset |
| Total number included in analysis (remission) | 158 | 101 | 169 | 110 | 221 | 159 | 326 | 1244 |
| Number relapsed at 6-8 months (%) | 32<br>(20) | 28<br>(27.7) | 19<br>(11.2) | 24<br>(21.8) | 34<br>(15.4) | 17<br>(10.7) | 107<br>(32.8) | 261<br>(21) |
| Number with one or more previous episodes of depression (%) | 112<br>(70.8) | 82<br>(81.9) | 104<br>(61.5) | 89<br>(80.9) | 154<br>(69.7) | 159<br>(100) | 124<br>(38) | 824<br>(66.2) |
| Mean PHQ-9 at baseline (SD) | 17.35<br>(4.20) | 15.33<br>(3.39) | 17.27<br>(4.05) | 16<br>(3.70) | 15.96<br>(3.92) | 15.77<br>(3.54) | 15.67<br>(4.17) | 16.17<br>(3.98) |
| Mean PHQ-9 at remission (SD) | 4.70<br>(2.69) | 5.19<br>(2.50) | 4.08<br>(2.56) | 5.97<br>(2.30) | 4.50<br>(2.61) | 4.53<br>(2.62) | 3.52<br>(2.31) | 4.40<br>(2.60) |
| Mean GAD-7 (SD) | 12.52<br>(4.83) | 8.25<br>(4.85) | 12.41<br>(4.96) | 11.80<br>(4.44) | GAD-7 not used in REEACT (CIS-R anxiety measure**) | 12.97<br>(4.44) | 13.62<br>(4.63) | 12.43<br>(4.92) |
| Mean age (SD) | 43.2 (12.9) | 71.8<br>(5.16) | 45.3<br>(13.9) | Not applicable (age is categorical*) | 40.3<br>(13.1) | 43.3<br>(14.7) | 41.7<br>(13.8) | 45.1<br>(15.63) |
| Gender:<br>Number<br>Female (%) | 113<br>(71.5) | 65<br>(64.3) | 100<br>(59.2) | 80<br>(72.7) | 148<br>(67) | 102<br>(64.2) | 190<br>(58.3) | 798<br>(64.1) |
| Ethnicity*:<br>Number White<br>(%) | 137<br>(86.7) | 100<br>(99) | 165<br>(97.6) | 107<br>(97.2) | 216<br>(97.7) | 154<br>(97.5) | 292<br>(94.2) | 1171<br>(95.4) |
| Employment*:<br>Number<br>Employed (%) | 133<br>(82.8) | Not<br>collected | 132<br>(78.1) | 105<br>(95.4) | 213<br>(96.3) | 149<br>(93.7) | 214<br>(65.6) | 946<br>(82.9) |
| Relationship<br>status*:<br>Number In a<br>relationship (%) | 69<br>(43.7) | Not<br>collected | 106<br>(62.7) | Not<br>collected | 140<br>(75.7) | 93<br>(58.9) | Not<br>collected | 408<br>(60.9) |
| Multimorbidity*:<br>Number with<br>multimorbidity<br>(%) | 88<br>(55.7) | 86<br>(85.1) | 96<br>(56.8) | Not<br>collected | Not collected | Not<br>collected | 87<br>(29.7) | 357<br>(49.5) |
| Current<br>antidepressant<br>use: Number<br>on<br>antidepressants<br>at remission<br>(%) | 113<br>(71.5) | 34<br>(41) | 128<br>(75.7) | 98<br>(90.7) | Not collected | 55<br>(41.4) | 130<br>(40.2) | 558<br>(57.3) |

\*See Supplemental Table 2.2 for further detail on categorical predictors
\*\*See Supplemental Table 2.3 for summary statistics for CIS-R anxiety subscale in REEACT

### 4.2. Univariable analysis

Table 2 presents the results from univariable multilevel models. Residual symptoms [OR: 1.13 (1.07-1.20)] and severity [OR: 1.07 (1.04-1.11)] were statistically significantly associated with relapse; number of previous episodes and comorbid anxiety were not.

**Table 2:**
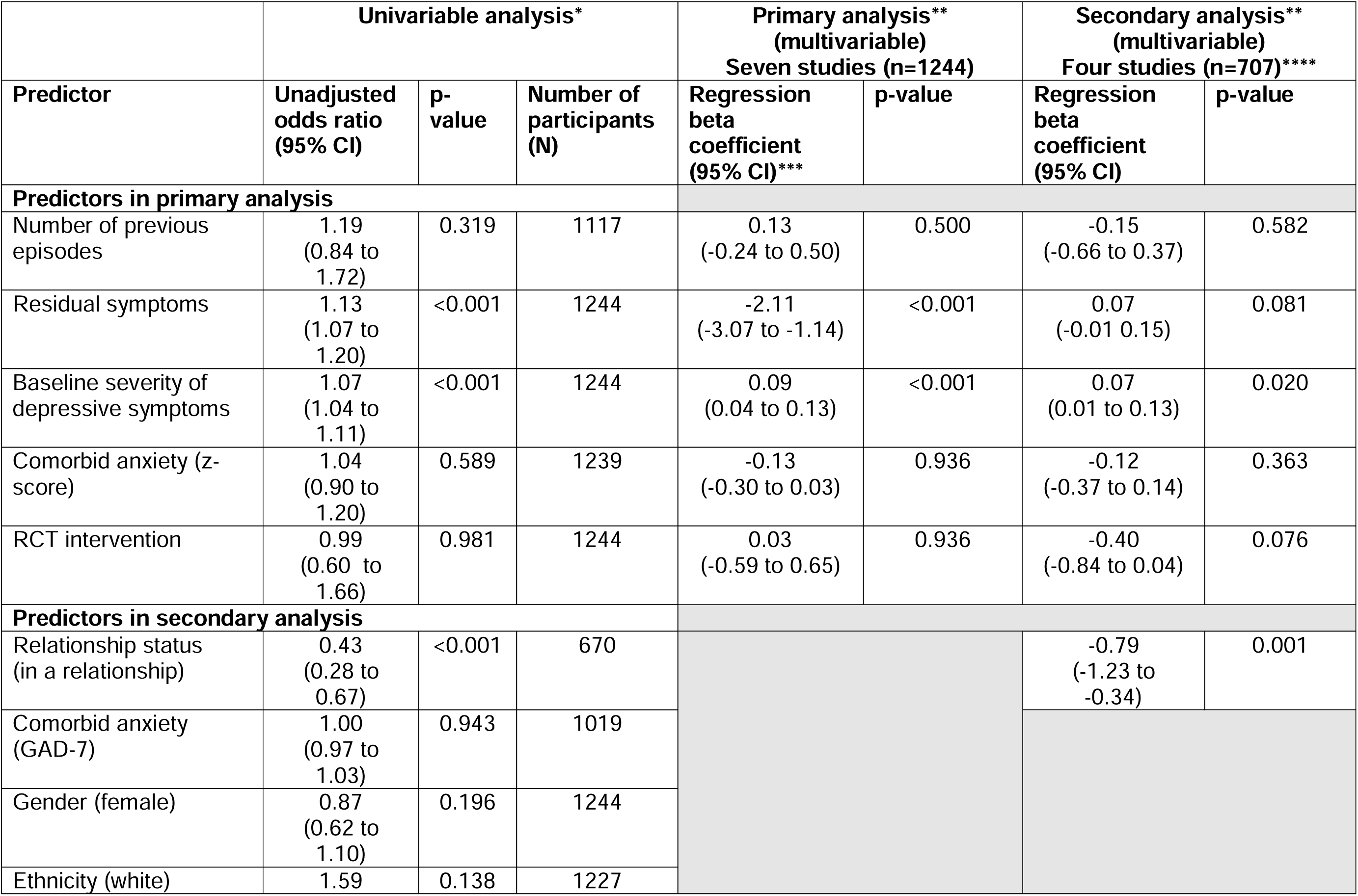
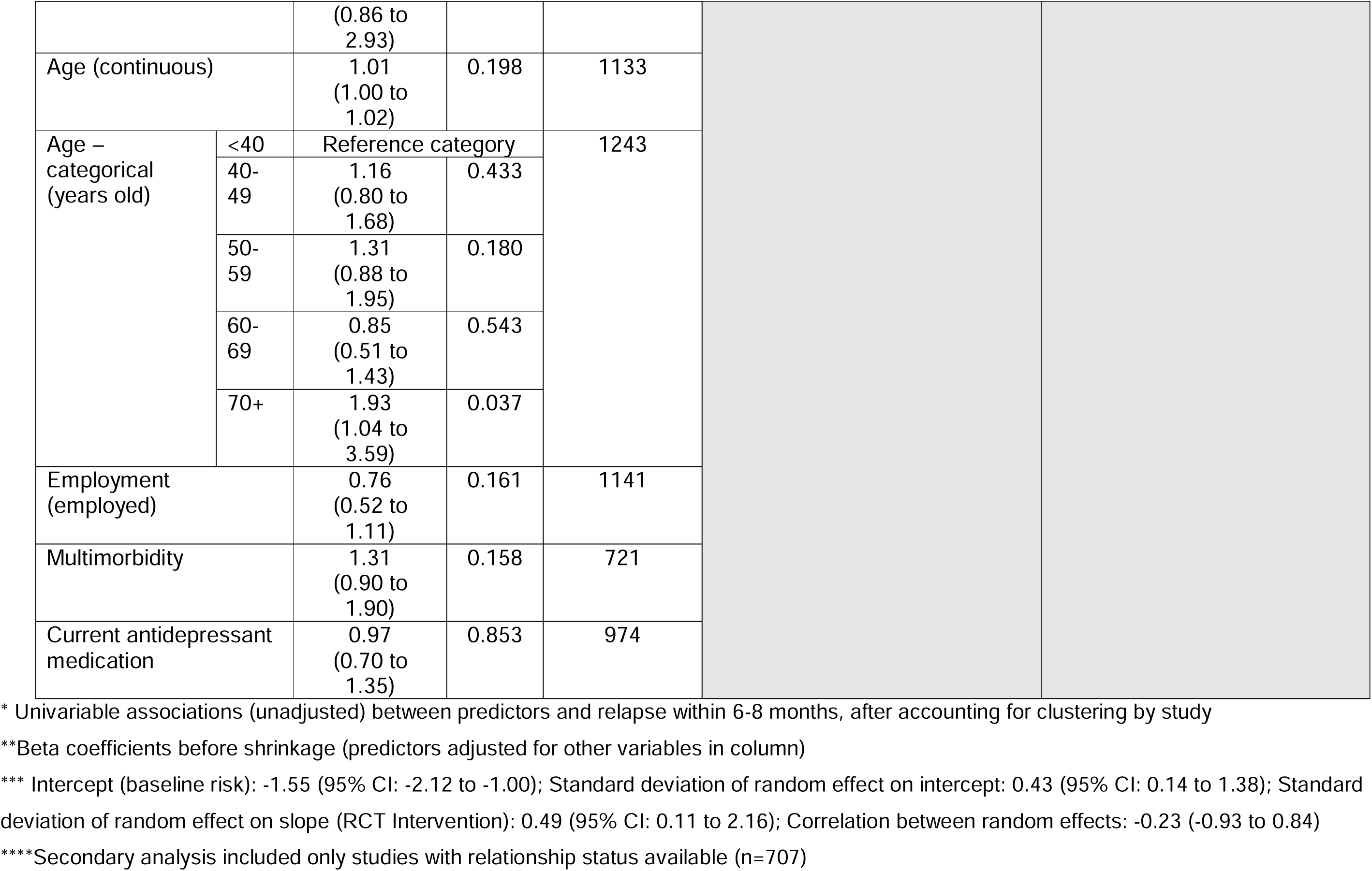
Univariable and multivariable associations between outcome and predictors (primary and secondary analysis)

|  | Univariable analysis* |  |  | Primary analysis**<br>(multivariable)<br>Seven studies (n=1244) |  | Secondary analysis**<br>(multivariable)<br>Four studies (n=707)**** |  |
| --- | --- | --- | --- | --- | --- | --- | --- |
| Predictor | Unadjusted odds ratio<br>(95% CI) | p-value | Number of participants<br>(N) | Regression beta coefficient<br>(95% CI)*** | p-value | Regression beta coefficient<br>(95% CI) | p-value |
| **Predictors in primary analysis** |  |  |  |  |  |  |  |
| Number of previous episodes | 1.19<br>(0.84 to 1.72) | 0.319 | 1117 | 0.13<br>(-0.24 to 0.50) | 0.500 | -0.15<br>(-0.66 to 0.37) | 0.582 |
| Residual symptoms | 1.13<br>(1.07 to 1.20) | <0.001 | 1244 | -2.11<br>(-3.07 to -1.14) | <0.001 | 0.07<br>(-0.01 to 0.15) | 0.081 |
| Baseline severity of depressive symptoms | 1.07<br>(1.04 to 1.11) | <0.001 | 1244 | 0.09<br>(0.04 to 0.13) | <0.001 | 0.07<br>(0.01 to 0.13) | 0.020 |
| Comorbid anxiety (z-score) | 1.04<br>(0.90 to 1.20) | 0.589 | 1239 | -0.13<br>(-0.30 to 0.03) | 0.936 | -0.12<br>(-0.37 to 0.14) | 0.363 |
| RCT intervention | 0.99<br>(0.60 to 1.66) | 0.981 | 1244 | 0.03<br>(-0.59 to 0.65) | 0.936 | -0.40<br>(-0.84 to 0.04) | 0.076 |
| **Predictors in secondary analysis** |  |  |  |  |  |  |  |
| Relationship status (in a relationship) | 0.43<br>(0.28 to 0.67) | <0.001 | 670 |  |  | -0.79<br>(-1.23 to -0.34) | 0.001 |
| Comorbid anxiety (GAD-7) | 1.00<br>(0.97 to 1.03) | 0.943 | 1019 |  |  |  |  |
| Gender (female) | 0.87<br>(0.62 to 1.10) | 0.196 | 1244 |  |  |  |  |
| Ethnicity (white) | 1.59 | 0.138 | 1227 |  |  |  |  |

|  |  |  |  |  |
| --- | --- | --- | --- | --- |
|  |  | (0.86 to 2.93) |  |  |
| Age (continuous) |  | 1.01<br>(1.00 to 1.02) | 0.198 | 1133 |
| Age – categorical (years old) | <40 | Reference category |  | 1243 |
|  | 40-49 | 1.16<br>(0.80 to 1.68) | 0.433 |  |
|  | 50-59 | 1.31<br>(0.88 to 1.95) | 0.180 |  |
|  | 60-69 | 0.85<br>(0.51 to 1.43) | 0.543 |  |
|  | 70+ | 1.93<br>(1.04 to 3.59) | 0.037 |  |
| Employment (employed) |  | 0.76<br>(0.52 to 1.11) | 0.161 | 1141 |
| Multimorbidity |  | 1.31<br>(0.90 to 1.90) | 0.158 | 721 |
| Current antidepressant medication |  | 0.97<br>(0.70 to 1.35) | 0.853 | 974 |

\* Univariable associations (unadjusted) between predictors and relapse within 6-8 months, after accounting for clustering by study
\*\*Beta coefficients before shrinkage (predictors adjusted for other variables in column)
\*\*\* Intercept (baseline risk): -1.55 (95% CI: -2.12 to -1.00); Standard deviation of random effect on intercept: 0.43 (95% CI: 0.14 to 1.38); Standard deviation of random effect on slope (RCT Intervention): 0.49 (95% CI: 0.11 to 2.16); Correlation between random effects: -0.23 (-0.93 to 0.84)
\*\*\*\*Secondary analysis included only studies with relationship status available (n=707)

### 4.3. Model development and apparent predictive performance

Table 2 presents the results of multivariable, multilevel logistic regression analysis for the primary analysis. The developed model, prior to shrinkage, had a pooled apparent performance of: C-statistic 0.62 (95% CI: 0.57-0.67), calibration slope of 0.95 (95% CI: 0.54-1.36), and calibration-in-the-large of 0.03 (95% CI: −0.49-0.54). See Supplemental Materials 3 for within-study performance statistics

### 4.4. Internal validation, shrinkage and final equation

Optimism-adjusted performance statistics, after bootstrapping, were: C-statistic 0.60, calibration slope 0.81, and calibration-in-the-large 0.03. The final model (Table 3) was produced by multiplying the original beta regression coefficients (from Table 2) by 0.81 (the optimism-adjusted calibration slope), and re-estimating the intercept to ensure calibration-in-the-large.

**Table 3:**
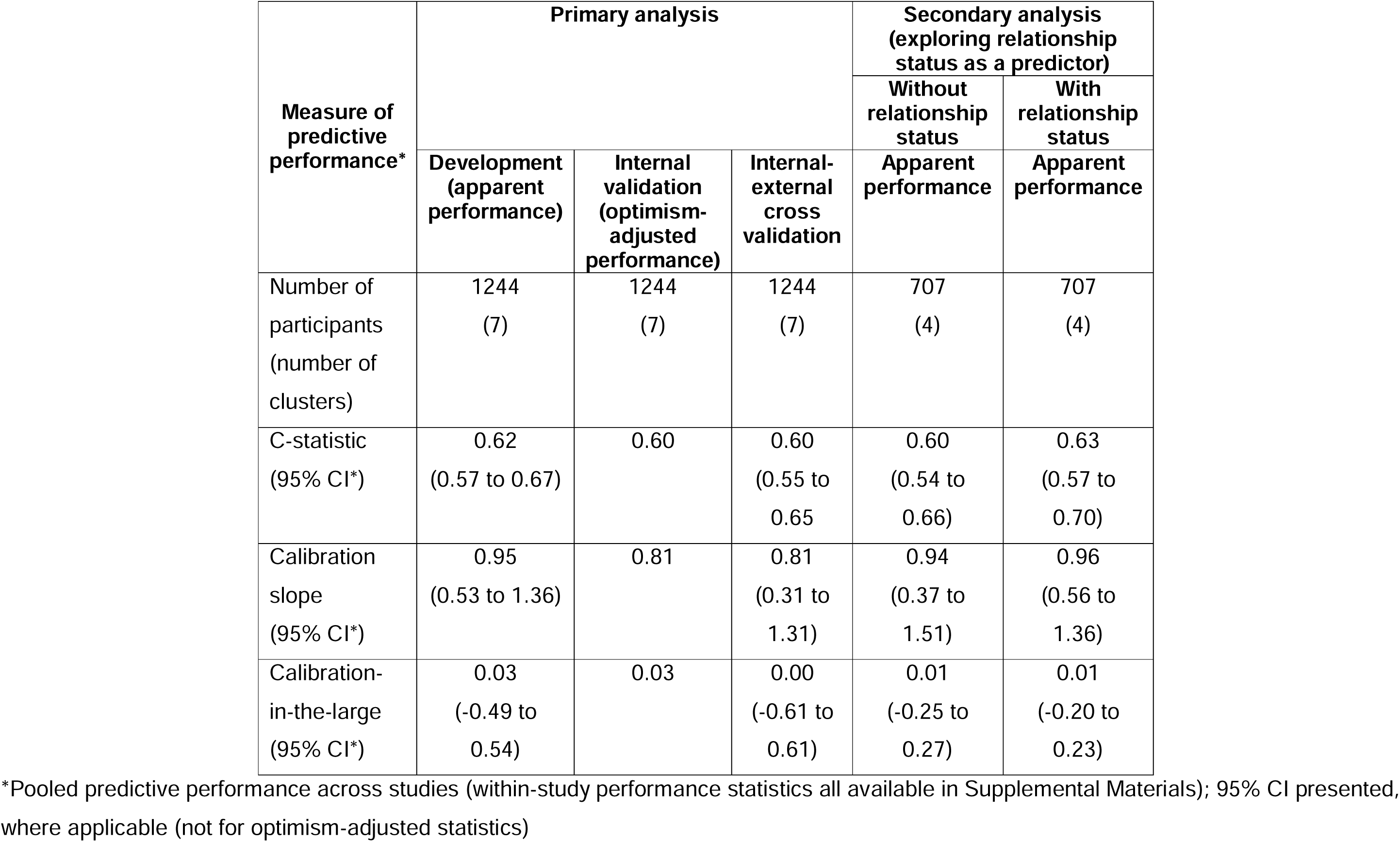
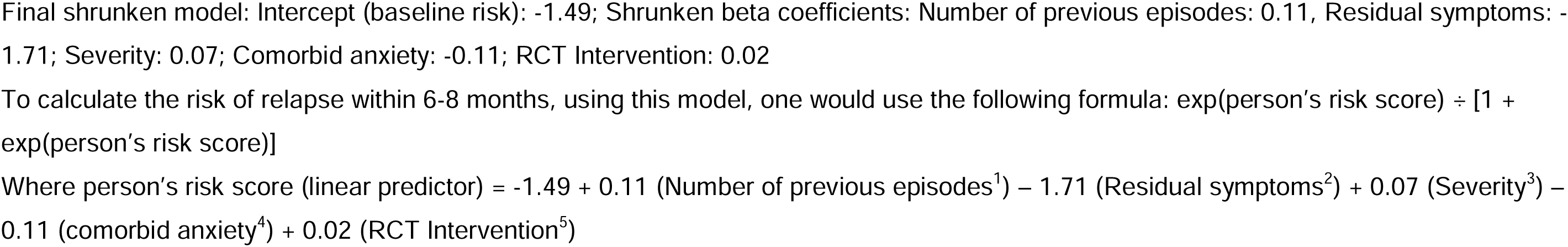
Summary of model’s predictive performance for primary, sensitivity and secondary analyses.

### 4.5. Internal-external cross-validation

Generalisability of the model was assessed using IECV(21). Calibration plots were compared for each validation in each of the different studies (Figure 1). These demonstrate inadequate calibration in most studies and significant heterogeneity in predictive performance across clusters. For example, WYLOW study shows severe miscalibration, with estimated risks generally too low, whereas in the COBRA study estimated risks are generally too high. In some studies, calibration was generally excellent (e.g., Healthlines Depression)

**Figure 1:**
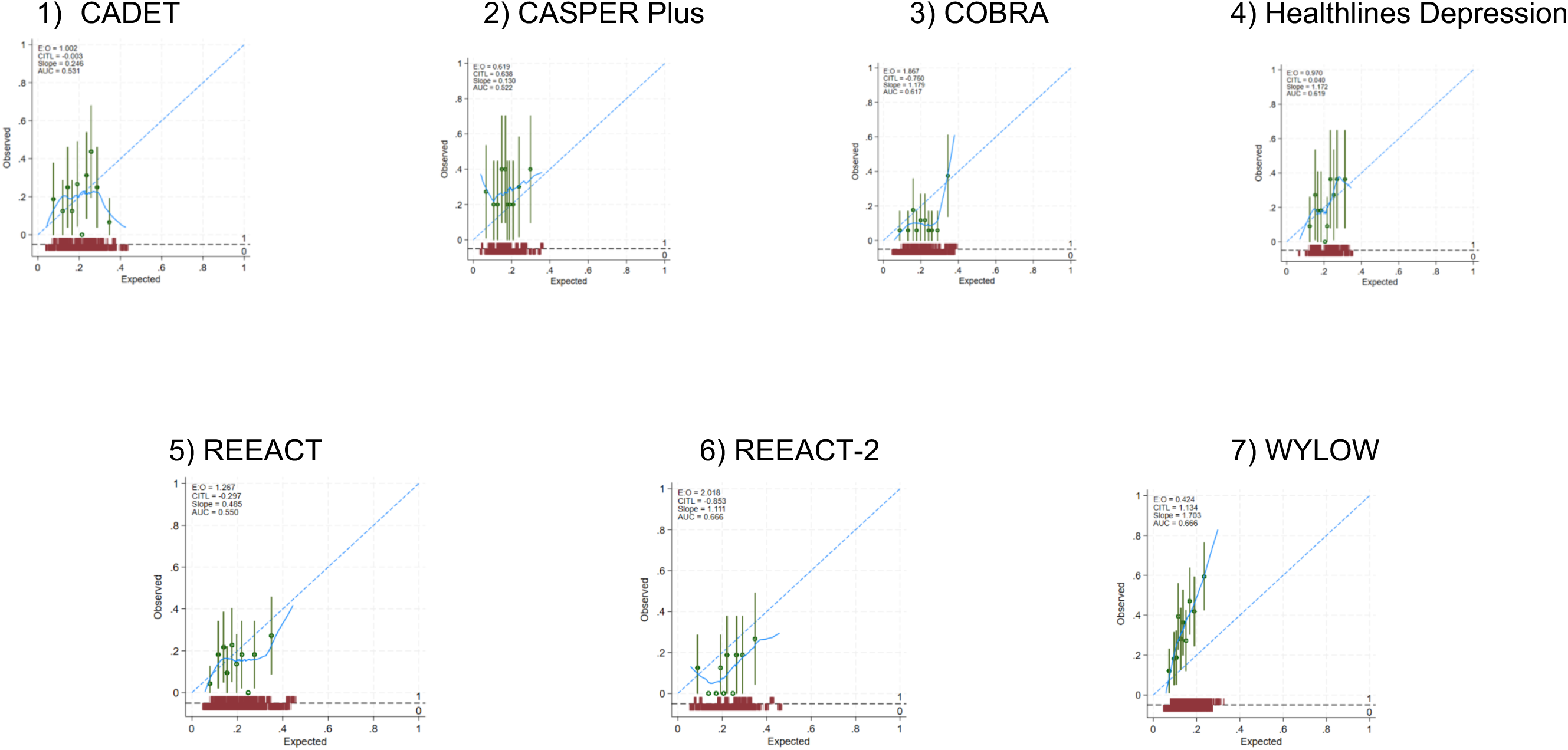
Calibration plots for internal-external cross-validation within each study

### 4.6. Sensitivity analysis

We removed REEACT and repeated the modelling process on the remaining six studies, to assess the impact of using z-scores to model comorbid anxiety. This did not change the study conclusions (see Supplemental Material 5 for analysis).

### 4.7. Secondary analysis

On univariable multilevel logistic analysis, relationship status was a highly statistically significant predictor (after adjusting the significance level to account for multiple significance testing using the Bonferroni correction). To further explore relationship status as a predictor of relapse, we repeated the model development procedures used in the primary analysis for the studies that included relationship status (CADET, COBRA, REEACT, and REEACT-2). We conducted these analyses both with and without the relationship status variable to provide a direct comparison (see Supplemental Material 6). Relationship status remained a statistically significant relapse predictor after adjusting for other prognostic factors (previous episodes, residual symptoms, severity, and comorbid anxiety).

## 5. DISCUSSION

We developed a model for predicting depression relapse in adults with remitted depression in primary care. Generally, the model had suboptimal predictive performance, with heterogeneous calibration across clusters on IECV and C-statistic below that required for acceptable discrimination. We would not recommend implementation in its current form, though calibration was promising in a subset of studies. Secondary analysis found a statistically significant association between relationship status and relapse.

### 5.1. Findings in the context of the literature

The performance of this model was in line with the performance measures for previous relapse prediction models(8). Residual symptoms were associated with relapse, which is consistent with existing literature(5,14). Residual symptoms are also associated with a more chronic depression course and poorer psychosocial functioning(22) and, as such, are an established treatment target in depression. The pre-existing evidence for severity as a prognostic factor for relapse is more equivocal than for residual symptoms(5). Residual symptoms are more likely in people with more severe initial depressive illness(23) and so the presence of residual symptoms may be a mediator of the relationship between baseline severity and relapse.

The lack of association between previous episodes and relapse in our study is not consistent with the consensus view(5,14). It is possible that this is because it was limited to a binary variable (“no previous episodes” or “one or more previous episodes”) in our study. However, the prognostic effect of previous episodes on relapse is known to be strongest when comparing any number of previous episodes to no previous episodes rather than being related to a specific number of previous episodes(5). An alternative explanation may be that previous episodes is most strongly associated with recurrence (which occurs over a longer time period than relapse) and therefore our 6-8 month follow-up was not sufficient to detect this association(5). Comorbid anxiety is recognised as a predictor of relapse(5); in particular, higher levels of anxiety at baseline have been found to predict a shorter time to relapse after treatment(24). In this study, we used anxiety at baseline (when depressed), using GAD-7 and the CIS-R anxiety subscale as measures of comorbid anxiety. It may be that an isolated measure of anxiety symptom severity at a single time-point is a crude measure and less important than knowing an individual’s history of comorbid anxiety.

Marital status (being single) is a risk factor for developing depression(17). A recent study also identified marital status (being single or no longer married) as being associated with a worse prognosis (more depressive symptoms) at 3-4 months (but not beyond 3-4 months)(25). While marital status is not an established predictor of relapse(5,17), our systematic review of prognostic models identified low-quality evidence of an association between relapse and marital status(8). The lack of association between relapse and other exploratory predictors (age, gender, ethnicity, employment status and multimorbidity) is consistent with other findings from the literature(5,14).

The prognostic factor research cited here is based on longitudinal studies, which examine the predictive value of specific variables based on sample-level trends. As our systematic review (8) and the current study demonstrate, combining these well-evidenced relapse predictors to produce individualised predictions of relapse risk remains challenging and calibration remains suboptimal for the purpose of personalised decision-making(7).

### 5.2. Strengths and limitations

This study was conducted according to best practice recommendations for methodology and reporting, with a sufficient sample size to produce precise risk estimates(26). By pre-selecting predictors with a robust, pre-existing evidence base, we aimed to mitigate the risk of overfitting and additional uncertainty associated with more data-driven approaches for predictor selection. While cohort studies and RCTs are recommended sources of data for prognostic model development(7), participants may differ from the general population in important ways and results should be interpreted with this in mind. For example, the majority of participants in this study were white, which limited our ability to explore ethnicity as a predictor. Some potentially useful predictors were not included (e.g., neuroticism, childhood maltreatment and rumination(5)) as they were not coded for in our cohorts (these are also not routinely measured in general practice settings). There was a risk of selection bias given the way studies were selected for inclusion. There was heterogeneity in IPD study populations (e.g., CASPER Plus included older adults), settings and treatment dose. WYLOW followed-up patients after low-intensity CBT whereas interventions in other studies (like COBRA) were more intensive and delivered over a greater number of sessions. While interventions were controlled for in the analysis, this heterogeneity could explain some of the observed miscalibration. Finally, outcomes (remission and relapse) were defined according to PHQ-9 (which is less optimal than diagnostic interview) and over a time-period (6-8 months) necessitated by the IPD data collection points [although one that is aligned with established definitions of relapse(5)].

### 5.3. Implications for future research

The strong statistically significant association between relationship status and relapse in our study warrants further confirmatory prognostic factor research going forwards, to better understand the association between marital/relationship and relapse and how this relates to its better understood association with depression more generally. Further research is needed to better understand whether other relapse predictors can be captured and recorded in an acceptable and valid way by primary care health professionals. For example, we know that routinely asking people about childhood maltreatment is not harmful(27), therefore it may be feasible to explore the use of childhood maltreatment and adversity as part of routine relapse risk assessment in primary care. There have been some efforts to develop clinically useful and valid brief instruments to measure rumination(28), which could be explored in a primary care setting. If there were robust evidence supporting the clinical utility of measuring and documenting additional relapse predictors, health professionals might then adopt this as routine practice. Better data linkage and systems integration across health and wellbeing services may also help address this problem.

In this study, we modelled the outcome of relapse as a binary outcome, which enabled us to incorporate criteria for reliable and significant change to define the outcome. It would be informative as part of future work to model the outcome on its continuous scale (i.e., build a linear model that estimates the outcome value, rather than the probability of a binary outcome), with dichotomisation done post-prediction. We focused on outcome occurrence by 6-8 months, but other time-points may be of interest.

As the IECV in this study demonstrated, there was heterogeneity in the external performance and generalisability of the model was not guaranteed. The studies from which IPD were drawn were generally comparable in terms of baseline demographic variables such as age, gender and ethnicity. Our IECV used the mean intercept and predictor-outcome associations to estimate performance. An alternative approach that could be considered in future work is local recalibration or intercept selection, where similarities in the outcome frequency or baseline characteristics (for example, mean age or proportion female) of a new population of interest is used to guide the intercept when applying the model in a different context(29). If the predictive performance of relapse prediction models can be improved in the future through recalibration or updating, clinical usefulness (using net benefit analysis) must be considered prior to implementation.

## 6. CLINICAL IMPLICATIONS

We cannot currently predict an individual’s risk of relapse with a high degree of accuracy, and existing relapse risk prediction models are unlikely to be suitable to guide the provision of relapse prevention. There are different approaches to prevention: universal approaches, which target whole populations; selective approaches, which target higher-risk groups; and indicated approaches, directed at individuals. In the absence of sufficiently accurate relapse risk prediction tools, we argue that a universal approach to relapse prevention of depression in primary care is currently warranted. This is likely to require a systems approach to mitigating the risk of and improving the management of relapse for all patients. This could mean targeting treatment at known prognostic factors (for example, focussing on reducing residual symptoms) or providing interventions as part of treatment during the acute phase of depression that target mechanisms of relapse (for example, prioritising relapse prevention planning)(30). Longer term, brief, inexpensive and scalable relapse prevention interventions are likely to be required for use in primary care.

## Data Availability

All data produced in the present study are available upon reasonable request to the authors.

## ACKNOWLEDGEMENTS

Thank you to the patient advisory group without whose contributions this study would not have been possible: Joanne Castleton, Penney Mayall, Gillian Payne, Sue Penn, and Emma Williams.

## AUTHOR CONTRIBUTIONS

ASM, CCG, SG, SA and DM conceptualised the study and acquired funding. ASM led on data curation, study administration and oversight, and wrote the original draft. ASM, LWP, KIES, LA, RDR, JEJB and NM contributed to methodology and data analysis. SG, JD, DAR and CS were responsible for collecting the data and agreed to share these for the purpose of this study. All authors contributed to the pre-registered protocol and study plan and all authors contributed to and approved the final manuscript.

## Footnotes

1 No previous episodes = 0; One or more previous episodes = 1

2 X^ −0.5 - 0.43 (where X = (residual_symptoms+1)) – this is the adjustment for non-linear transformation and mean-centring

3 Severity – 16.17

4 Comorbid_anx_zscore + 0.118

5 This would be zero when applied in clinical practice, outside the context of an RCT

