## Supplemental Material for "The development and validation of a prognostic model to predict relapse in adults with remitted depression in primary care: secondary analysis of pooled individual participant data from multiple studies"

Table 1.1: Sources of individual participant data

| Study | N | Study type | Inclusion criteria | Length of Follow-up | Follow up Points | Mean age (SD) | Gender (% Female) | RCT Intervention | Duration of RCT Intervention |
|---|---|---|---|---|---|---|---|---|---|
| CADET | 581 | RCT | Adults with depression | 12 months | 0, 4,12 | 44.4 (13.3) | 71.9 | Collaborative care | 14 weeks |
| CASPER Plus | 485 (358 at 12 m) | RCT | 65 years or older with depression | 18 months | 0, 4, 12 and 18 months | Intervention group: 71.9 (6.03)<br><br>Control: 71.6 (5.96) | Intervention group: 59.1<br><br>Control: 63.1 | Collaborative care | 8-10 weeks |
| COBRA | 440 | RCT | Adults with depression | 18 months | 0, 6, 12, 18 | 43.5 (14.1) | 66 | Behavioural Activation vs CBT | 16 weeks |
| Healthlines Depression | 609 | RCT | Adults with depression | 12 months | 0, 4,8,12 | Intervention group: 49.1 (12.9)<br><br>Control: 50 (12.8) | Intervention group: 69<br>Control: 68 | Complex intervention (Integrated telehealth) | 12 months |

| | | | | | | | | | |
|---|---|---|---|---|---|---|---|---|---|
| REEACT | 461 | RCT | Adults with depression | 24 months | 0, 4, 12 and 24 months | 39.86 (12.65) | 67 | cCBT | 6 weeks |
| REEACT-2 | 369 | RCT | Adults with depression | 12 months. | 0, 4 and 12 months | 40.6 (13.8) | 64.5 | cCBT | 4 months |
| WYLOW | 439 | Longitudinal observational cohort study | Adults with depression | 12 months (Start-point = Remission) | Monthly | 41.28 (14.59) | 59.7 | None (cohort were followed up after LiCBT) | NA |

Table 1.2: CIS-R anxiety subscale

| Item | Description | Score |
|---|---|---|
| Compulsions | A series of questions around compulsions | +1 if repeating actions >3 days in past week<br>+1 if attempted to stop<br>+1 if action is upsetting<br>+1 if an action was repeated more than twice |
| Anxiety | A series of questions around anxiety | +1 if anxious for >3days in past week<br>+1 if causes feeling of unpleasantness<br>+1 if causes physical symptoms<br>+1 if anxious >3hrs in any day |
| Irritability | A series of questions around feelings of irritability, anger or short-temper | +1 if consistent during the past week (>3 days)<br>+1 if feeling lasted >1hr in any day during past week<br>+1 if shouted or felt like shouting<br>+1 if lost temper without reason |
| Worry | A series of questions around worry | +1 if worry persists for >3days in past week<br>+1 if excessively worried<br>+1 if worry was unpleasant<br>+1 if worried >3hrs in any day |
| Panic | A series of questions around panic | +1 if panic occurred once in past week<br>+1 if panic occurred at least once more in past week<br>+1 if panic attack lasted longer than 10 mins<br>+1 if panic attack was unpleasant |
| Phobias | A series of questions around phobias | +1 if anxious for >3days in past week<br>+1 if causes physical symptoms<br>+1 if avoidance action taken for at least 1day<br>+1 if avoidance action taken for more than 3days |
| Obsessions | A series of questions around the presence of obsessive thoughts | +1 if consistent during the past week (>3 days)<br>+1 if tried to stop<br>+1 if they are upsetting<br>+1 lasted for at least 15 mins |
| Health anxiety | A series of questions about concern over health or future health | +1 if consistent during the past week (>3 days)<br>+1 if considered excessive<br>+1 if considered unpleasant<br>+1 if difficult to stop worrying |
| Somatic concerns | A series of questions around the presence | +1 if consistent during the past week (>3 days). |

| | | |
|---|---|---|
| | of aches/pains or bodily discomfort | +1 if consistent and lasted at least 3 in any day during last week<br>+1 if consistent and unpleasant<br>+1 if consistent and bothersome during 'interesting' activity |

Table 1.3: Re-categorisation of categorical variables for analysis

| Variable | Original categories (in RCTs) | New categories (PREDICTR) |
|---|---|---|
| Ethnicity | White | White |
| | Mixed | Other |
| | Black | |
| | Asian | |
| | Chinese | |
| | Other | |
| Employment status | Employed (Full time or part time) | Employed/not seeking employment |
| | Student | |
| | Retired | |
| | House-person | |
| | Unemployed due to ill-health | |
| | Other | |
| | Unemployed and seeking work | Unemployed |
| Relationship status | Married/civil partnership/cohabiting/relationship | In a relationship |
| | Single | Single |
| | Separated | |
| | Divorced | |
| | Widowed | |
| Multimorbidity | None | No long-term physical health condition |
| | Mental health only | |
| | Diabetes | One or more long-term physical health conditions |
| | Asthma or COPD | |
| | Degenerative or inflammatory arthritis | |
| | Heart Disease | |
| | Stroke | |
| | Cancer | |

Table 1.4: Master codebook for IPD Harmonisation

| Code | Description of code | Type of data | Method of measurement | Range of values and coding of predictors |
|---|---|---|---|---|
| **ID_orig** | ID in original study | Identifier (individual participant) | NA | NA |
| **ID_PREDICTR** | ID assigned for purpose of PREDICTR IPD dataset | Identifier (individual participant) | NA | NA |
| **RCT** | Coding for individual studies | Identifier (cluster) | NA | CADET=1<br>CASPER Plus=2<br>COBRA=3<br>Healthlines=4<br>REEACT=5<br>REEACT-2=6<br>WYLOW=7 |
| **PHQ_baseline** | PHQ-9 at Baseline of RCT | Continuous | PHQ-9 | 0-27 |
| **GAD_baseline** | GAD-7 at baseline | Continuous | GAD-7 | 0-21 |
| **PHQ_FU1** | t=0 for PREDICTR | Continuous | PHQ-9 | 0-27 |

| | | | | |
|---|---|---|---|---|
| **PHQ_FU2** | 6-8 months post-FU1 | Continuous | PHQ-9 | 0-27 |
| **followup_time_(FU2-FU1)** | How long has elapsed between FU1 and FU2 | NA | NA | 6 or 8 (months) |
| **remit** | Has the pt remitted at FU1?<br>If PHQ-9 score at baseline > 10 and less than 10 at FU1 (plus change of 5 or more points) | Binary | NA | Yes=1, No=0 |
| **relapse** | Has the pt relapsed at FU2?<br>If remission at FU1 and PHQ-9 >10 (plus change of 5 or more points) | Binary | NA | Yes=1, No=0 |
| **residual_symptoms (=PHQ_FU1)** | PHQ-9 at remission | Continuous | PHQ-9 score at remission | 0-9 |

| | | | | |
|---|---|---|---|---|
| **prev_eps** | Number of previous episodes of depression | Categorical | Patient or GP report (No previous episodes vs any previous episodes) | No previous episodes=0; 1 or more previous episodes=1 |
| **comorbid_anx (=GAD_baseline)** | Comorbid anxiety | Continuous | GAD-7 Score | 0-21 |
| **comorbid_anx_zscore** | Comorbid anxiety (z score) | Continuous | Combined z score based on mean and SD within original study dataset<br><br>GAD-7 for all studies other than REEACT<br><br>For REEACT, z score of CIS-R anxiety subscale | NA |
| **Severity (=PHQ_baseline)** | Severity of depression at baseline | Continuous | PHQ-9 score at baseline | 10-27 |
| **RCT_intervention** | Presence or absence of | Categorical | Presence or absence of treatment | Absence of effective treatment |

| | | | | |
|---|---|---|---|---|
| | RCT intervention | | | (control arm or non-effective intervention)=0; Presence of treatment (effective intervention arm)=1 |
| **age** | Age in years | Continuous | Self-report | |
| **gender** | Gender | Categorical | Self-report | Male=0, Female=1 |
| **ethnicity** | Ethnicity | Categorical | Self-report | White=0 Other=1 |
| **employment** | Employment status | Categorical | Self-report | Unemployed=0; Employed=1 |
| **relationship** | Relationship status | Categorical | Self-report | In a relationship=1; Not in a relationship=0 |
| **multi_morbidity** | Multi-morbidity | Categorical | Self-report | No long-term physical health condition=0; One or more long-term physical health conditions=1 |
| **ADM_current** | Current antidepressant | Categorical | Self-report | Not currently taking ADM at remission=0; |

| | | | | |
|---|---|---|---|---|
| | medication (ADM) | | | Taking ADM at remission=1 |

Figure 1.1: Flow diagram of participants in PREDICTR dataset

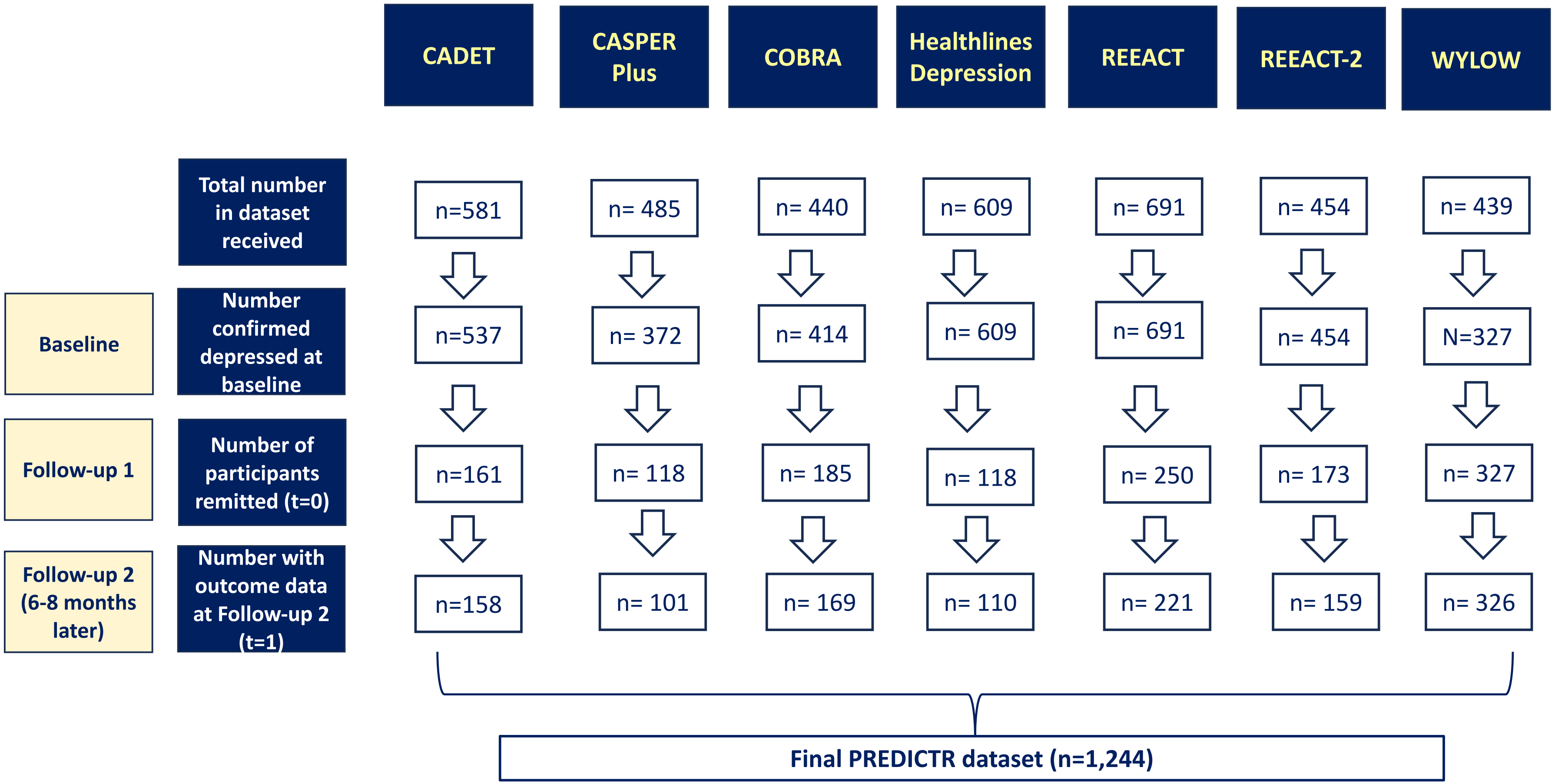

#### 1.1. Changes from protocol

The full protocol and statistical analysis plan was pre-registered and published. Following inspection of the data, some changes to the pre-registered analysis plan were necessary, outlined here.

1. The protocol included a further (eighth) study [COINCIDE (45)] for inclusion in the PREDICTR dataset. The IPD from COINCIDE could not be used in the analysis as there was data only from baseline, 4 months and 24 months and, as a result, we could not define the outcome of relapse within 6-8 months for this study.

2. The IPD from REEACT did not use the GAD-7 to measure anxiety. Instead, the authors had used the Clinical Interview Schedule-Revised (CIS-R). Rather than discard the IPD from REEACT, we made the decision to use the anxiety subscale from the CIS-R as a measure of comorbid anxiety. We converted this to standardised scores (z-scores) and used this to model this predictor, along with z-scores for GAD-7 from the other six studies. Z-scores were calculated within each cluster for the whole dataset, prior to removing those who had not reached remission. For the primary analysis, therefore, comorbid anxiety was measured as z-score rather than GAD-7 as planned. To test the validity of this, we conducted a sensitivity analysis removing REEACT from the analysis and using GAD-7 to assess the impact of this decision.

3. Given the number of systematically missing exploratory predictors (see Supplemental Table 5), the pre-planned exploratory analysis, using data-driven predictor selection, could not be performed. However, we were able to measure univariable associations between these exploratory predictors and relapse. Where this association was statistically significant, we measured the effect on predictive performance of including the predictor in the model. The Bonferroni method was used to account for multiple significance testing to

provide adjusted p-values for use as thresholds for significance. This was done to reduce the risk of false positive significant associations after multiple testing during the exploratory analysis.

4. The planned sensitivity analysis omitting WYLOW and COBRA were not deemed necessary as the IECV included a development analysis omitting WYLOW and COBRA (and used these as validation sets).

5. The definition of “unemployed” changed from the protocol, prior to any analysis. We adapted the categorisation of employed to include those unemployed but not seeking work due to ill health.

### 2. Descriptive statistics and risk of bias assessment

Table 2.1: Availability of variables and missing data in individual participant data

| Variable | | Variable present in study | | | | | | | Total number with predictor* | Total number missing (%) |
|---|---|---|---|---|---|---|---|---|---|---|
| | | CADET (n=158) | CASPER Plus (n=101) | COBRA (n=169) | Healthlines Depression (n=110) | REEACT (n=221) | REEACT-2 (n=159) | WYLOW (n=326) | | |
| Previous episodes | Available in study? | ✓ | ✓ | ✓ | ✓ | ✓ | ✓ | ✓ | 1244 | 127 (10.2) |
| | Number of participants with missing data (%) | None | None | 16 (9.5) | 13 (11.8) | None | None | 98 (30) | | |
| Residual symptoms | Available in study? | ✓ | ✓ | ✓ | ✓ | ✓ | ✓ | ✓ | 1244 | 0 |
| | Number of participants with missing data (%) | None | None | None | None | None | None | None | | |
| Severity | Available in study? | ✓ | ✓ | ✓ | ✓ | ✓ | ✓ | ✓ | 1244 | 0 |
| | Number of participants with missing data (%) | None | None | None | None | None | None | None | | |
| Comorbid anxiety: GAD-7 | Available in study? | ✓ | ✓ | ✓ | ✓ | | ✓ | ✓ | 1023 | 4 (0.4) |
| | Number of participants with | 2 (1.3) | 1 (1) | None | 1 (0.9) | | None | None | | |

| | | | | | | | | | | |
|---|---|---|---|---|---|---|---|---|---|---|
| | missing data (%) | | | | | | | | | |
| Comorbid anxiety: CIS-R (% missing) | Available in study? | | | | | ✓ | | | 221 | 1 (0.5) |
| | Number of participants with missing data (%) | | | | | 1 (0.5) | | | | |
| Age-continuous (% missing) | Available in study? | ✓ | ✓ | ✓ | | ✓ | ✓ | ✓ | 1134 | 1 (0.09) |
| | Number of participants with missing data (%) | None | None | None | | 1 (0.5) | None | None | | |
| Age-categorical | Available in study? | | | | ✓ | | | | 110 | 0 |
| | Number of participants with missing data (%) | | | | None | | | | | |
| Gender | Available in study? | ✓ | ✓ | ✓ | ✓ | ✓ | ✓ | ✓ | 1244 | 0 |
| | Number of participants with missing data (%) | None | None | None | None | None | None | None | | |
| Ethnicity | Available in study? | ✓ | ✓ | ✓ | ✓ | ✓ | ✓ | ✓ | 1244 | 17 (1.4) |
| | Number of participants with | None | None | None | None | None | 1 (0.6) | 16 (4.9) | | |

| | | | | | | | | | | |
|---|---|---|---|---|---|---|---|---|---|---|
| | missing data (%) | | | | | | | | | |
| Employment status | Available in study? | ✓ | | ✓ | ✓ | ✓ | ✓ | ✓ | 1143 | 2 (0.2) |
| | Number of participants with missing data (%) | 1 (0.6) | | None | 1 (0.9) | None | None | None | | |
| Relationship status | Available in study? | ✓ | | ✓ | | ✓ | ✓ | | 707 | 37 (5.2) |
| | Number of participants with missing data (%) | None | | None | | 36 (16.3) | 1 (0.6) | | | |
| Multimorbidity | Available in study? | ✓ | ✓ | ✓ | | | | ✓ | 754 | 33 (4.4) |
| | Number of participants with missing data (%) | None | None | None | | | | 33 (10.1) | | |
| Antidepressant use at remission | Available in study? | ✓ | ✓ | ✓ | ✓ | | ✓ | ✓ | 1023 | 49 (4.8) |
| | Number of participants with missing data (%) | None | 18 (17.8) | None | 2 (1.8) | | 26 (16.4) | 3 (0.9) | | |

✓ = variable present in study; *Number of participants in the combined studies with predictor available (including those with missing data)

Table 2.2: Summary of categorical predictors and their coding in original datasets

| Predictor variable | Study (total number of participants in study) | | | | | | | |
|---|---|---|---|---|---|---|---|---|
| | | **CADET (n=158)** | **CASPER Plus (n=101)** | **COBRA (n=169)** | **Healthlines Depression (n=110)** | **REEACT (n=221)** | **REEACT-2 (n=159)** | **WYLOW (n=326)** |
| **RCT Intervention** | Effective RCT Intervention | Collaborative care: n=87 | Collaborative care: n=56 | CBT: n=92<br><br>BA: n=77 | Healthlines Integrated Telehealth intervention: n=67 | RCT Intervention (cCBT) was not effective | MoodGym with telephone support: n=80 | Not applicable. All participants received LiCBT through IAPT. |
| | Ineffective RCT Intervention or Control | Usual care: n=72 | Usual care: n=45 | No usual care arm: n=0 | Usual care: n=43 | cCBT ("Beating the Blues"): n=66;<br>cCBT ("MoodGym"): n=78;<br>Usual care: n=77 | Guided self-help with telephone support: n=30;<br>MoodGym only: n=49 | |
| **Ethnicity** | White | White: n=137 | White: n=100 | White British: n=155;<br>White Irish: n=4;<br>White (other): n=6 | White: n=107 | White British: n=208;<br>Any other White background: n=8 | White British: n=145;<br>White Irish: n=3;<br>Any other White background: n=6 | White: n=292 |
| | Non-white | Asian or Asian British: n=8;<br>Black or Black British: n=7;<br>Mixed: n=3;<br>Other: n=3 | Black or Black British: n=1 | Other Asian: n=1;<br>Black African: n=1;<br>Other=1;<br>Prefer not to say: n=1 | Mixed: n=2;<br>Other: n=1 | Asian or Asian British: n=1;<br>Chinese: n=1;<br>Japanese: n=1;<br>Jewish: n=1 | Asian or Asian British (Indian): n=1<br>Chinese: n=1<br>Other (not specified): n=2 | Mixed: n=7;<br>Asian: n=5;<br>Black: n=4;<br>Chinese: n=1;<br>Other: n=2 |
| | Missing | Missing: n=0 | Missing: n=0 | Missing: n=0 | Missing: n=0 | Missing: n=0 | Missing: n=1 | Missing: n=16 |

| | | | | | | | | |
|---|---|---|---|---|---|---|---|---|
| **Relationship status** | In a relationship | Married/living as married: n=69 | | Cohabiting: n=19; Civil partnership: n=1; Married: n=86 | | Married: n=100; Living with a partner: n=38; In a relationship: n=2 | Married: n=71; Living with partner: n=22 | |
| | Not in a relationship | Single: n=49; Separated: n=13; Divorced: n=23; Widowed: n=4 | | Single: n=32; Divorced / separated: n=31 | | Divorced / separated: n=21; Widowed: n=2; Never married: n=22 | Divorced/separated: n=21; Widowed: n=4; Single: n=39; Other (not specified): n=1 | |
| | Missing | Missing: n=0 | | Missing: n=0 | | Missing: n=36 | Missing: n=1 | |
| **Employment status** | Employed or not seeking employment | Full-time paid or self employment: n=66; Part-time paid or self employment: n=29; Voluntary employment: n=1; Student: n=3; Housewife / husband: n=12; Retired: n=13; Other=9 | | Employed / student: n=120; Retired: n=12 | Full time work: n=35; Part time work: n=18; Full time education: n=1; Unable to work (illness): n=15; Unable to work (carer): n=1; Retired: n=20; Looking after home: n=7; Other: n=9 | Employed part-time: n=37; Employed full-time: n=92; Self=employed: n=21; Retired: n=14; Looking after family or home: n=9; Not employed (ill health): n=15; Not employed but not seeking: n=3; Other: n=22 | Employed part-time: n=26; Employed full-time: n=62; Self-employed: n=15; Retired: n=16; Looking after family or home: n=5; Not employed but not seeking work due to ill health: n=4; Full-time student: n=16; Other (job lined up): n=1 | Employed, retired, student, homemaker: n=214 |

| | | | | | | | | |
|---|---|---|---|---|---|---|---|---|
| | Unemployed | Unemployed: n=27 | | Not employed: n=37 | Unemployed: n=4 | Not employed but seeking work: n=8 | Not employed but seeking work: n=10 | Unemployed: n=112 |
| | Missing | Missing: n=0 | | Missing: n=0 | Missing: n=1 | Missing: n=0 | Missing: n=0 | Missing: n=0 |
| **Multimorbidity** | Multi-morbidity | Diabetes: n=2; Asthma: n=14; Arthritis: n=7; Heart disease: n=3; High blood pressure: n=11; More than one of the above: n=11; Other: n=40 | At least one of diabetes, osteoporosis, hypertension, rheumatoid arthritis, osteoarthritis, stroke, cancer, respiratory condition, eye condition, or heart disease: n=86 | One or more long-term conditions (in addition to depression): n=96 | | | | Long-term condition (self-reported): n=87 |
| | No multi-morbidity | No long-standing illness, disability or infirmity: n=70 | None of above: n=15 | None: n=73 | | | | No long-term condition (self-reported): n=206 |
| | Missing | Missing: n=0 | Missing: n=0 | Missing: n=0 | | | | Missing: n=33 |
| **Age (categorical)** | <40 years old | n=64 | n=0 | n=59 | n=22 | n=96 | n=68 | n=145 |
| | 40-49 years old | n=40 | n=0 | n=46 | n=25 | n=69 | n=39 | n=85 |
| | 50-59 years old | n=35 | n=0 | n=37 | n=32 | n=39 | n=24 | n=64 |
| | 60-69 years old | n=17 | n=43 | n=18 | n=22 | n=12 | n=25 | n=24 |

| | | | | | | | | |
|---|---|---|---|---|---|---|---|---|
| | 70 years and over | n=2 | n=58 | n=9 | n=9 | n=4 | n=3 | n=8 |

Table 2.3: Summary statistics for CIS-R anxiety subscale in REEACT

| Item | Mean score (SD)* | |
|---|---|---|
| | **REEACT Total (n=685)** | **REEACT PREDICTR (n=221)** |
| Compulsions | 0.72 (1.19) | 0.47 (0.97) |
| Anxiety | 2.19 (1.51) | 2.01 (1.55) |
| Irritability | 2.16 (1.36) | 1.94 (1.38) |
| Worry | 2.46 (1.33) | 2.38 (1.38) |
| Panic | 0.74 (1.25) | 0.44 (1.03) |
| Phobias | 1.34 (1.28) | 1.15 (1.15) |
| Obsessions | 1.26 (1.60) | 1.06 (1.49) |
| Health anxiety | 0.88 (1.15) | 0.70 (1.03) |
| Somatic concerns | 1.51 (1.44) | 1.33 (1.38) |
| Total | 13.27 (6.66) | 11.45 5.86) |

*For REEACT dataset as a whole (REEACT Total) and for those included in PREDICTR study (i.e. those who have remitted) (REEACT PREDICTR):

Table 2.4: Detailed risk of bias assessment (PROBAST) for sources of IPD

| | **Study** | | | | | | |
|---|---|---|---|---|---|---|---|
| | **CADET** (Richards et al., 2013) | **CASPER Plus** (Bosanquet et al., 2017) | **COBRA** (Richards et al., 2016) | **Healthlines Depression** (Salisbury et al., 2016) | **REEACT** (Gilbody et al., 2015) | **REEACT-2** (Gilbody et al., 2017) | **WYLOW** (Ali et al., 2017) |
| **Domain 1: Participants** | | | | | | | |
| 1.1. Appropriate data sources? | Yes | Yes | Yes | Yes | Yes | Yes | Yes |
| 1.2. Appropriate inclusions and exclusion? | Yes | Yes | Yes | Yes | Yes | Yes | Yes |
| **Risk of bias** | **Low** | **Low** | **Low** | **Low** | **Low** | **Low** | **Low** |
| **Domain 2: Predictors** | | | | | | | |
| 2.1. Defined and assessed in similar way for all participants? | Yes | Yes | Yes | Yes | Yes | Yes | Yes |
| 2.2. Assessments made without knowledge of outcome? | Yes | Yes | Yes | Yes | Yes | Yes | Yes |
| 2.3. All available at | Yes | Yes | Yes | Yes | Yes | Yes | Yes |

| | | | | | | | |
|---|---|---|---|---|---|---|---|
| time of model's intended use? | | | | | | | |
| **Risk of bias** | **Low** | **Low** | **Low** | **Low** | **Low** | **Low** | **Low** |
| **Domain 3: Outcome** | | | | | | | |
| 3.1. Determined appropriately? | Yes | Yes | Yes | Yes | Yes | Yes | Yes |
| 3.2. Pre-specified or standard definition? | Yes | Yes | Yes | Yes | Yes | Yes | Yes |
| 3.3. Predictors excluded from outcome definition? | Yes | Yes | Yes | Yes | Yes | Yes | Yes |
| 3.4. Defined and determined similar for all participants? | Yes | Yes | Yes | Yes | Yes | Yes | Yes |
| 3.5. Determined without knowledge of predictors? | Yes | Yes | Yes | Yes | Yes | Yes | Yes |
| 3.6. Appropriate time interval between predictor assessment | Yes | Yes | Yes | Yes | Yes | Yes | Yes |

| | | | | | | | |
|---|---|---|---|---|---|---|---|
| and outcome determination? | | | | | | | |
| **Risk of bias** | **Low** | **Low** | **Low** | **Low** | **Low** | **Low** | **Low** |
| **Overall assessment of risk of bias** | **Low** | **Low** | **Low** | **Low** | **Low** | **Low** | **Low** |

### 3. Model development and apparent performance

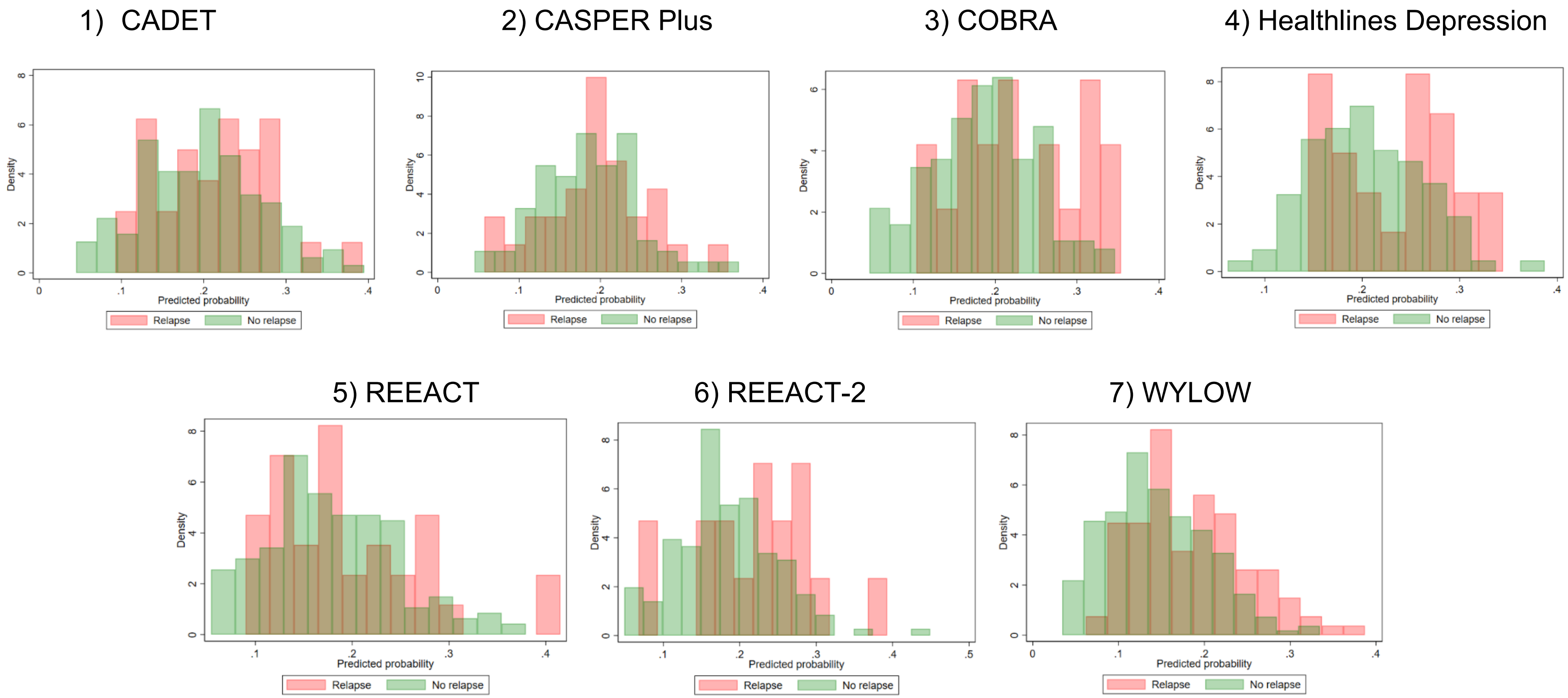

Figure 3.1: Distribution of predicted probabilities (apparent performance before shrinkage) by observed outcomes in each cluster

Table 3.2: Within-cluster and pooled (apparent) predictive performance statistics for primary analysis model

| Study | N total (N relapsed) | C-statistic (95% CI) | Calibration slope (95% CI) | Calibration-in-the-large (95% CI) |
|---|---|---|---|---|
| CADET | 158 (32) | 0.56 (0.45 to 0.67) | 0.50 (-0.34 to 1.33) | 0.01 (-0.38 to 0.41) |
| CASPER Plus | 101 (28) | 0.55 (0.42 to 0.68) | 0.25 (-0.73 to 1.24) | 0.53 (0.08 to 0.97) |
| COBRA | 169 (19) | 0.64 (0.49 to 0.79) | 1.45 (0.22 to 2.69) | -0.63 (-1.11 to -0.15) |
| Healthlines Depression | 110 (24) | 0.63 (0.50 to 0.76) | 1.35 (0.01 to 2.70) | 0.05 (-0.41 to 0.50) |
| REEACT | 221 (34) | 0.55 (0.45 to 0.66) | 0.56 (-0.23 to 1.34) | -0.21 (-0.58 to 0.17) |
| REEACT-2 | 159 (17) | 0.66 (0.51 to 0.81) | 1.12 (-0.08 to 2.33) | -0.67 (-1.18 to -0.16) |
| WYLOW | 326 (107) | 0.68 (0.62 to 0.74) | 1.48 (0.93 to 2.03) | 1.00 (0.76 to 1.23) |
| Pooled results | 1244 (261) | 0.62 (0.57 to 0.67) | 0.95 (0.54 to 1.36) | 0.03 (-0.49 to 0.54) |

Figure 3.1: Pooled performance statistics for model development (apparent performance)

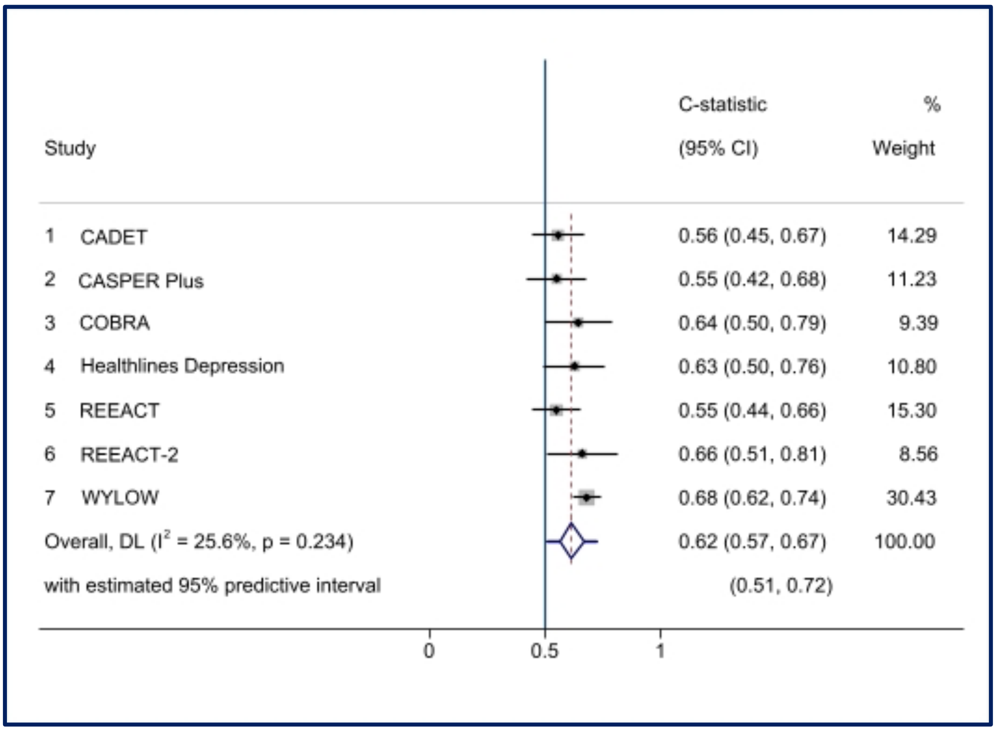

Figure 3.1(a): Forest plot showing within-cluster and pooled C-statistic (apparent performance)

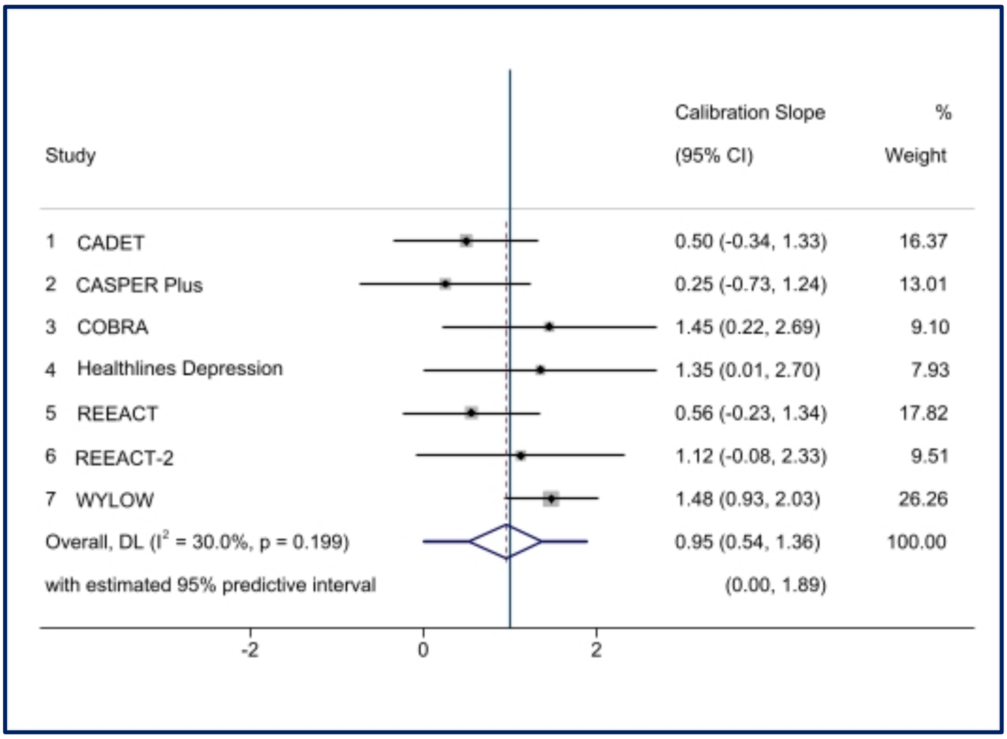

Figure 3.1(b): Forest plot showing within-cluster and pooled calibration slope (apparent performance)

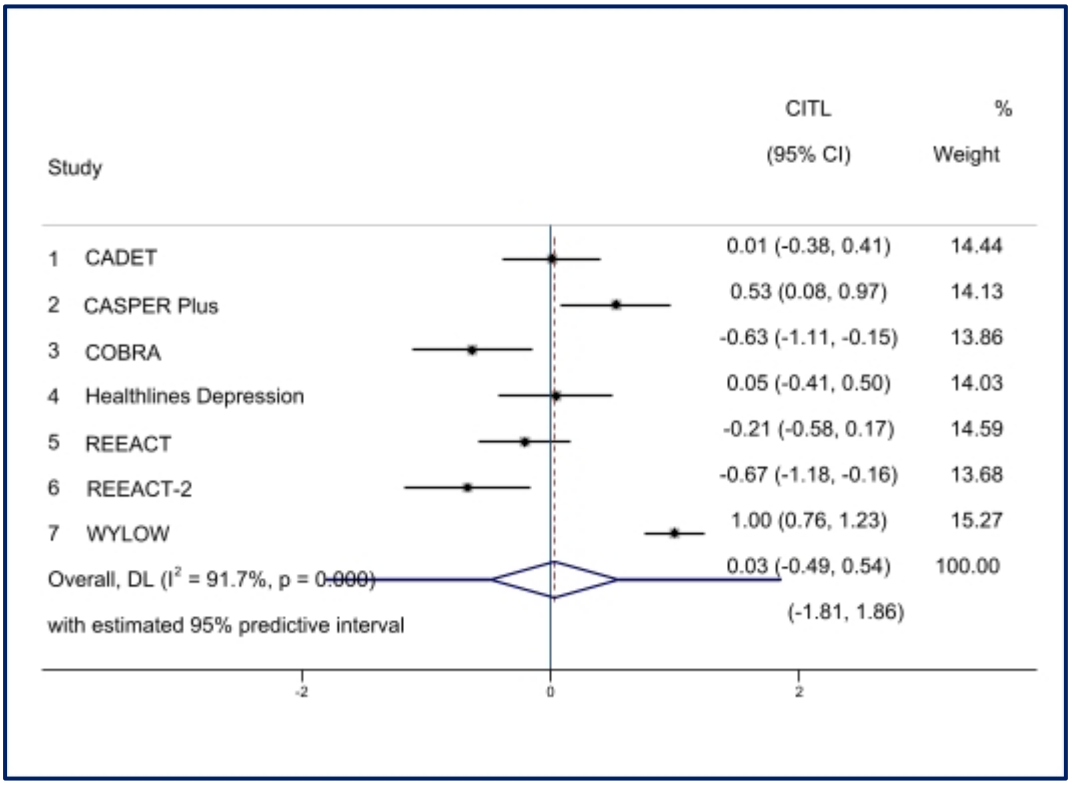

Figure 3.1(c): Forest plot showing within-cluster and pooled calibration-in-the-large (apparent performance)

Figure 3.2: Calibration plots (with calibration curves) showing apparent performance of developed model in each cluster

1) CADET

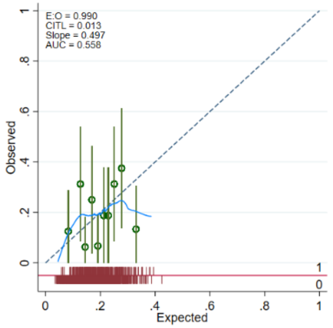

2) CASPER Plus

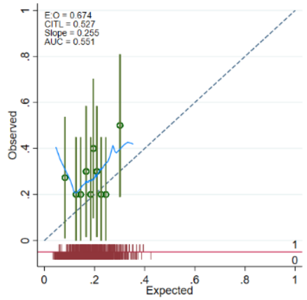

3) COBRA

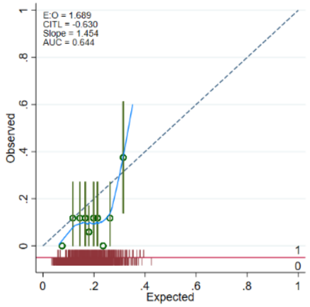

4) Healthlines Depression

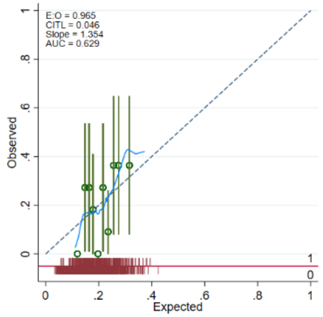

5) REEACT

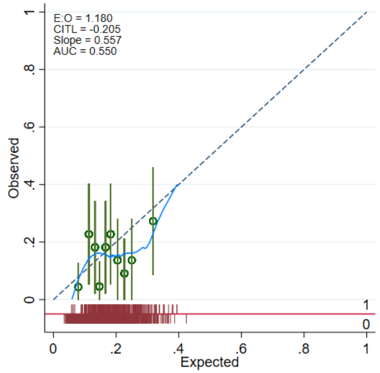

6) REEACT-2

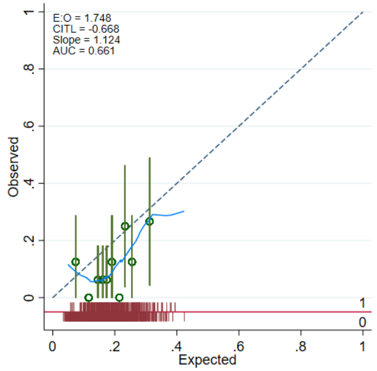

7) WYLOW

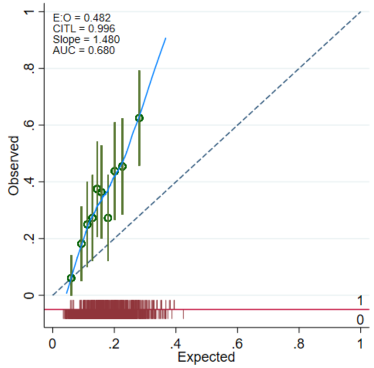

### 4. Internal-external cross validation (IECV)

**Full procedure:**

We excluded data from each primary study in turn and developed the risk prediction model in the remaining data, using the same model development approach as detailed (without shrinkage, as the purpose was primarily to explore the generalisability of the model). We then externally validated the developed model using the data from the excluded study. This process was repeated, each time omitting a different study, until the model had been fitted excluding each study once. Predictive performance metrics (C-statistic for discrimination; calibration slope, calibration-in-the-large and visual inspection of calibration plots with LOESS-smoothed calibration curves for calibration) were calculated for the final developed model in each "external" validation (that is, when the model was applied in the study that had been left out). Random effects meta-analysis was used to summarise the performance across studies, to obtain summary measures of the model performance and estimates of heterogeneity in performance across studies. 95% prediction intervals were also constructed to calculate the model's likely performance in new but similar settings.

Random effects meta-analyses were performed to summarise the performance statistics from each validation within each round of IECV. The pooled summary performance statistics are presented in Figure 4.1 and Table 4.1. The distributions of predicted probabilities for each cluster are displayed in Appendix 4.2. Calibration plots were compared for each validation in each of the different clusters (Figure 4.3). These demonstrate inadequate calibration and heterogeneity across clusters (for example, the calibration plot for REEACT-2 demonstrates over-prediction and the one for WYLOW shows under-prediction of outcome).

Figure 4.1: Forest plots showing pooled performance statistics for IECV

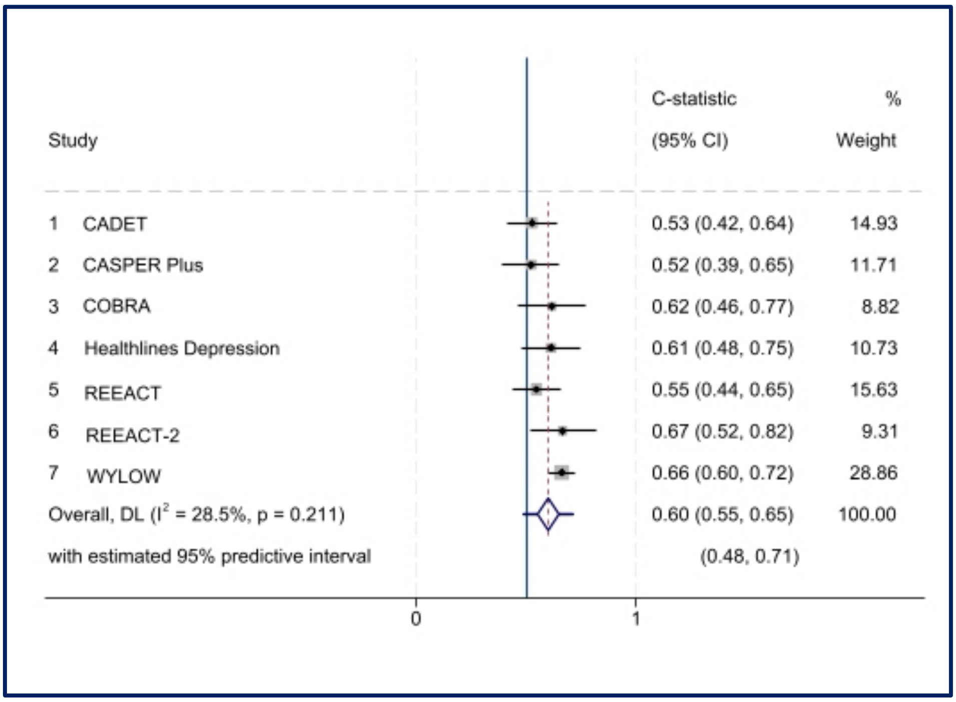

Figure 4.1(a): Forest plot showing C-statistic for each validation and pooled C-statistic in IECV

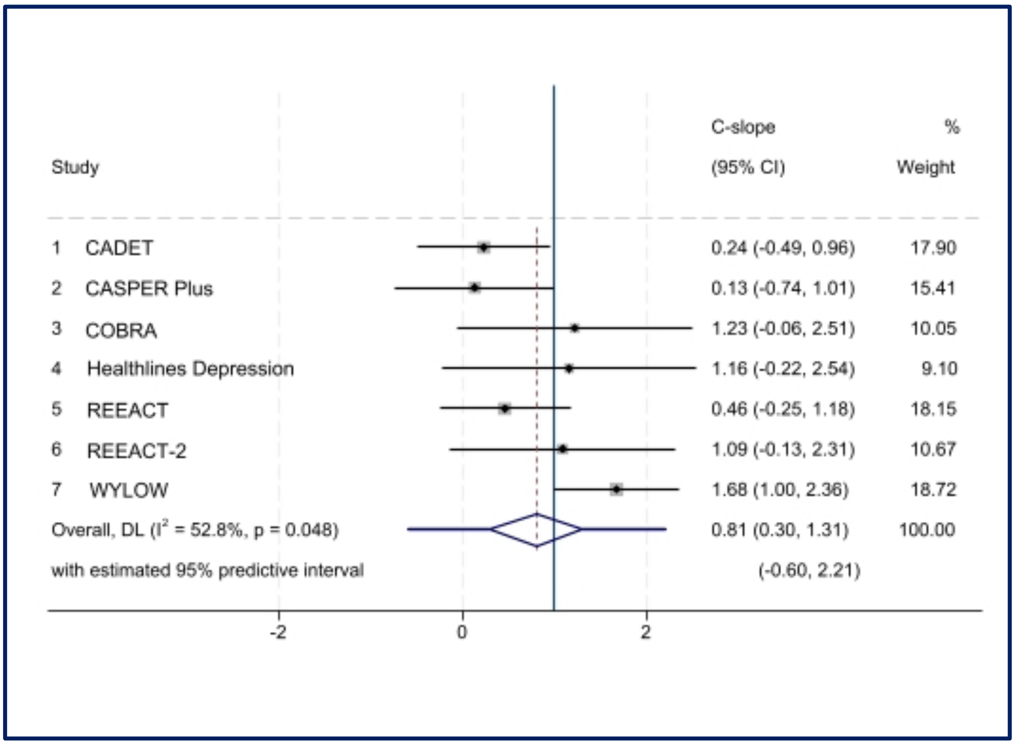

Figure 4.1(b): Forest plot showing calibration slope for each validation and pooled calibration slope in IECV

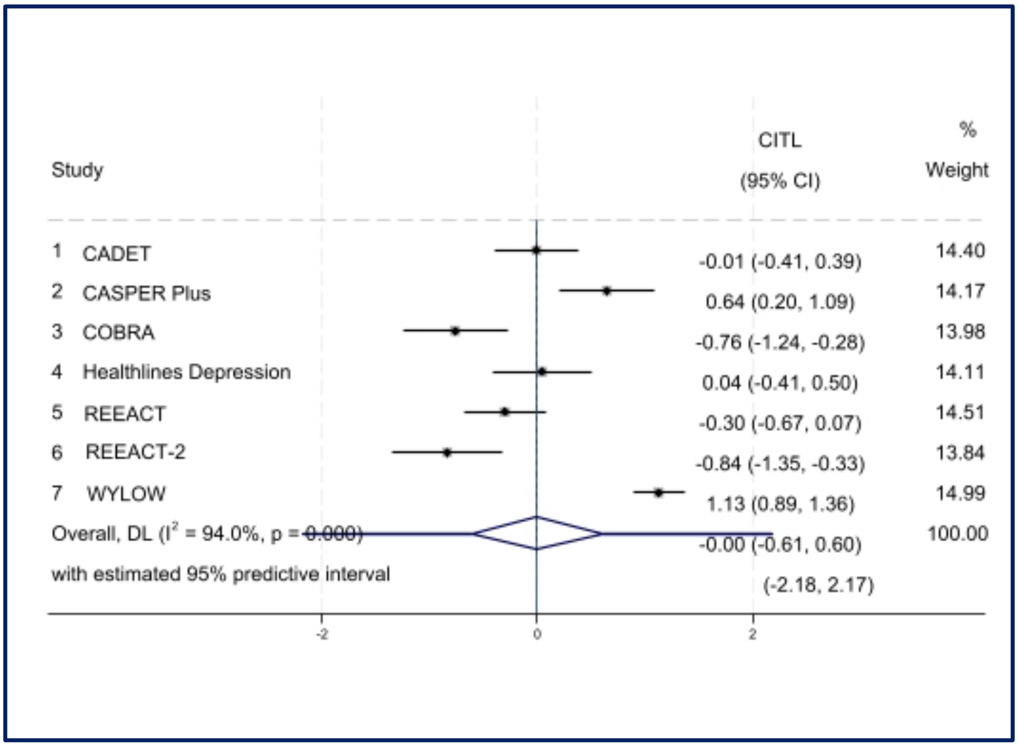

Figure 4.1(c): Forest plot showing CITL for each validation and pooled CITL in IECV

Table 4.1: Summary of performance statistics in each validation (IECV)

| **Study** | **N total (N relapsed)** | **C-statistic (95% CI)** | **Calibration slope (95% CI)** | **Calibration-in-the-large (95% CI)** |
|---|---|---|---|---|
| CADET | 158 (32) | 0.53 (0.42 to 0.64) | 0.24 (-0.49 to 0.96) | -0.01 (-0.41 to 0.39) |
| CASPER Plus | 101 (28) | 0.52 (0.39 to 0.65) | 0.13 (-0.74 to 1.01) | 0.64 (0.20 to 1.09) |
| COBRA | 169 (19) | 0.62 (0.47 to 0.77) | 1.23 (-0.06 to 2.51) | -0.76 (-1.24 to -0.28) |
| Healthlines Depression | 110 (24) | 0.61 (0.48 to 0.75) | 1.16 (-0.22 to 2.54) | 0.04 (-0.41 to 0.50) |
| REEACT | 221 (34) | 0.55 (0.44 to 0.65) | 0.46 (-0.25 to 1.18) | -0.30 (-0.67 to 0.07) |
| REEACT-2 | 159 (17) | 0.67 (0.52 to 0.82) | 1.09 (-0.13 to 2.31) | -0.84 (-1.35 to -0.33) |
| WYLOW | 326 (107) | 0.66 (0.60 to 0.72) | 1.68 (1.01 to 2.36) | 1.13 (0.89 to 1.36) |
| Pooled | 1244 (261) | 0.60 (0.55 to 0.65) | 0.81 (0.31 to 1.31) | 0.00 (-0.61 to 0.60) |

Figure 4.2: Predicted probability distributions in each cluster (IECV)

1) CADET

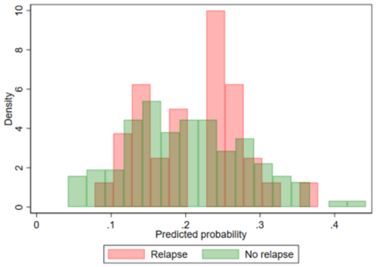

2) CASPER Plus

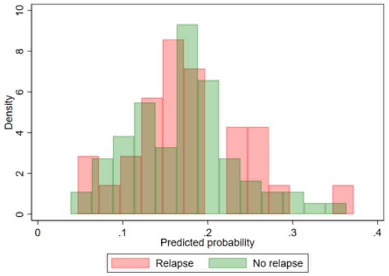

3) COBRA

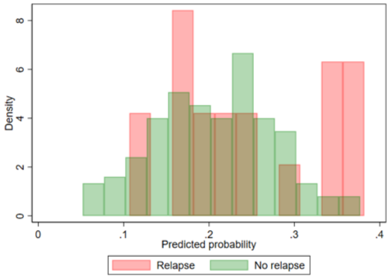

4) Healthlines Depression

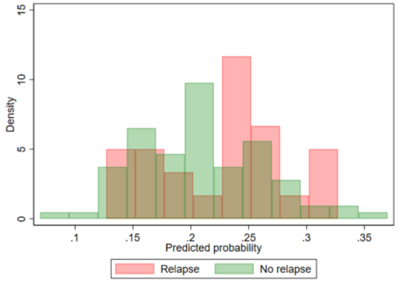

5) REEACT

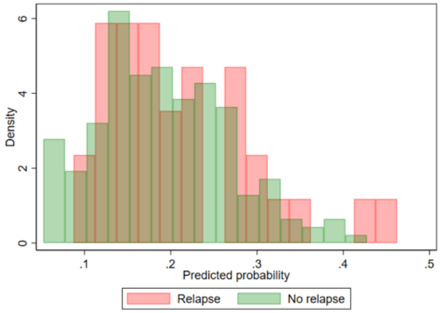

6) REEACT-2

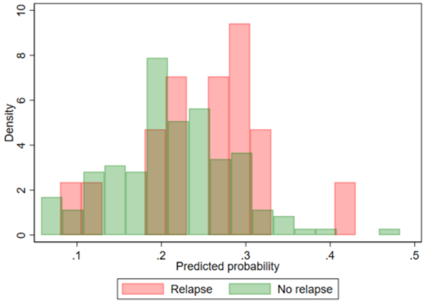

7) WYLOW

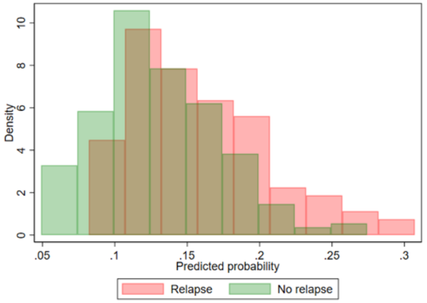

### 5. Sensitivity analysis

This sensitivity analysis excluded REEACT and modelled comorbid anxiety as GAD-7 rather than z-score in the other six studies. All other variables and modelling approach were the same as in the primary analysis.

*Modelling of continuous predictors*

MFPs were used to model continuous predictors and explore non-linear relationships within the imputed datasets, as for the primary analysis.

Table 5.1: Transformations and mean-centring of continuous predictors following MFP modelling

| Predictor | Transformation and centring |
|---|---|
| Residual symptoms | X^2-28.9576543<br>(X = (residual_symptoms+1)) |
| Residual symptoms 2 | X^2*ln(X)-48.73333691<br>(X = (residual_symptoms+1)) |
| Severity | severity-16.21515152 |
| Comorbid anxiety (GAD-7) | comorbid_anx-12.42807717 |

(Adjusted) results from the multivariable analysis are presented in Table 5.2.

Table 5.2: Results from multilevel multivariable associations (adjusted*) between outcome and predictors (sensitivity analysis)

| **Predictor** | **Beta coefficient (95% CI)** | **p-value** |
|---|---|---|
| Number of previous episodes | 0.08 (-0.35 to 0.51) | 0.715 |
| Residual symptoms | 0.15 (0.09 to 0.22) | <0.001 |
| Residual symptoms 2 | -0.06 (-0.09 to -0.03 | <0.001 |
| Severity | 0.10 (0.05 to 0.14) | <0.001 |
| Comorbid anxiety (GAD-7) | -0.04 (-0.08 to 0.00) | 0.047 |
| RCT intervention | 0.01 (-0.67 to 0.69) | 0.979 |

Intercept (baseline risk): -1.36 (95% CI: -2.06 to -0.67)

Standard deviation of random effect on intercept: 0.45 (95% CI: 0.12 to 1.72)

Standard deviation of random effect on slope (RCT Intervention): 0.54 (95% CI: 0.11 to 2.72)

Correlation between random effects: -0.32 (95% CI: -0.96 to 0.86)

*adjusted for other predictor variables within model

We calculated pooled performance statistics (C-statistic, C-slope and calibration-in-the-large) and also within-cluster statistics to assess heterogeneity in model apparent performance during model development. 95% Prediction intervals were also calculated.

Figure 5.1: Pooled performance statistics for sensitivity analysis

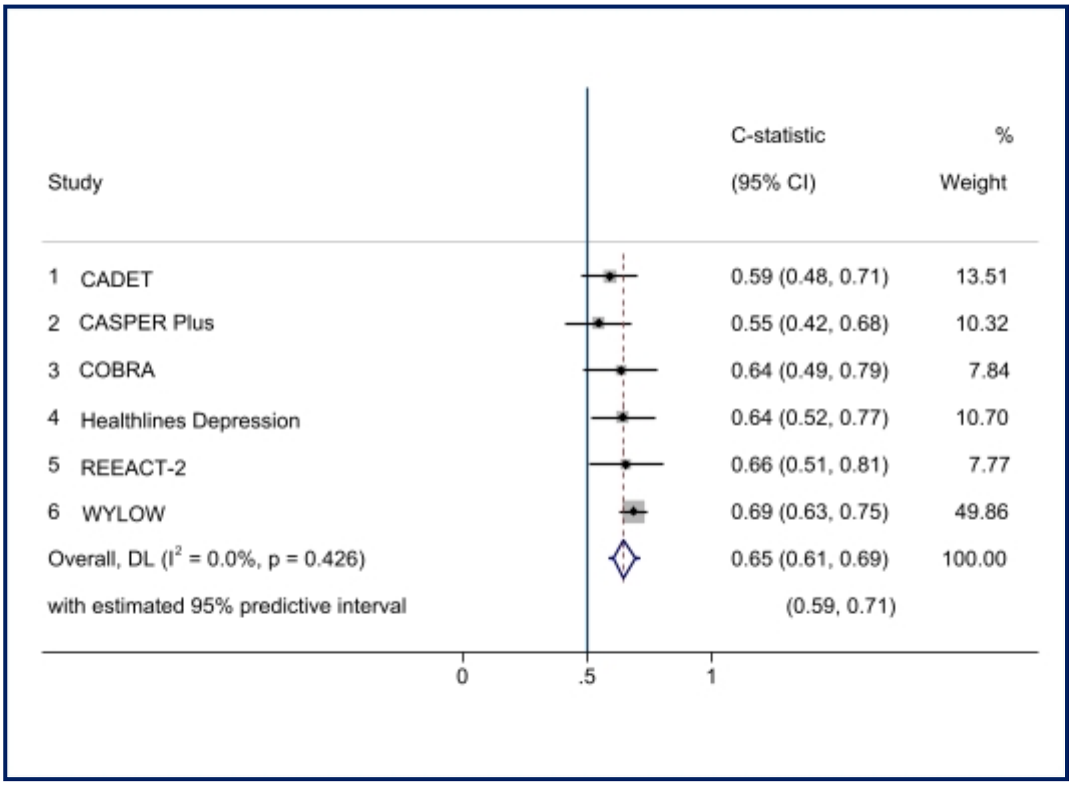

Figure 5.1(a): Pooled C-statistic for sensitivity analysis (apparent performance)

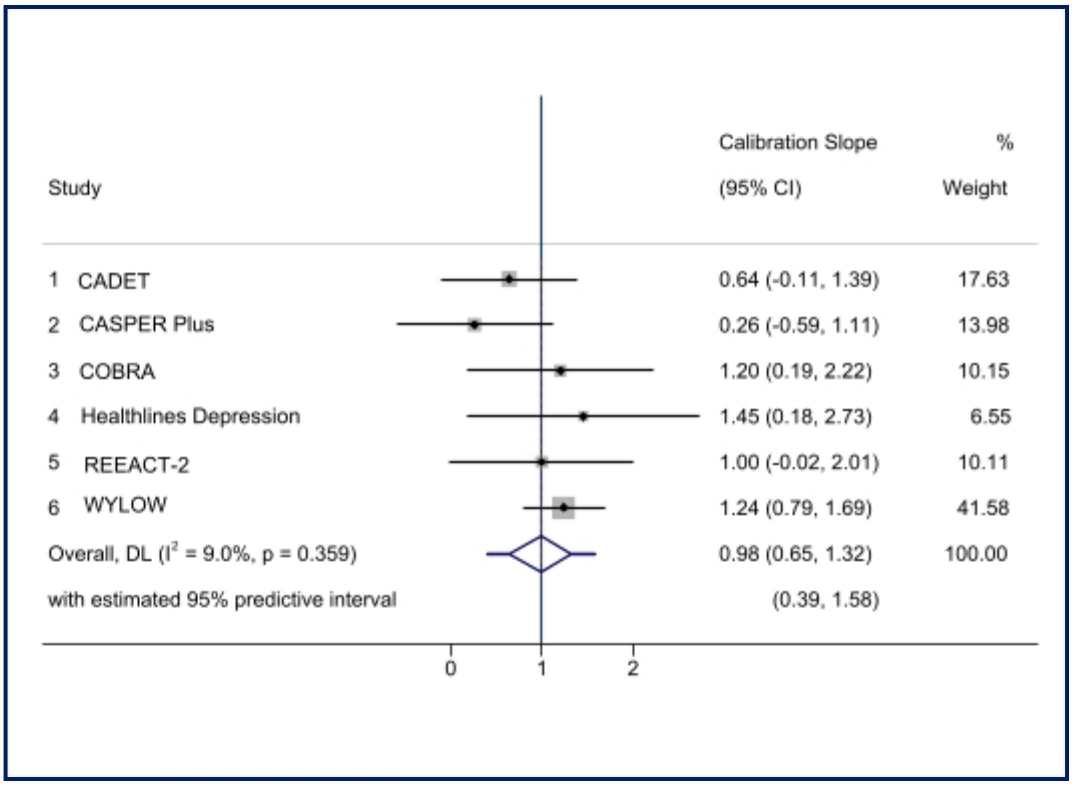

Figure 5.1(b): Pooled calibration slope for sensitivity analysis (apparent performance)

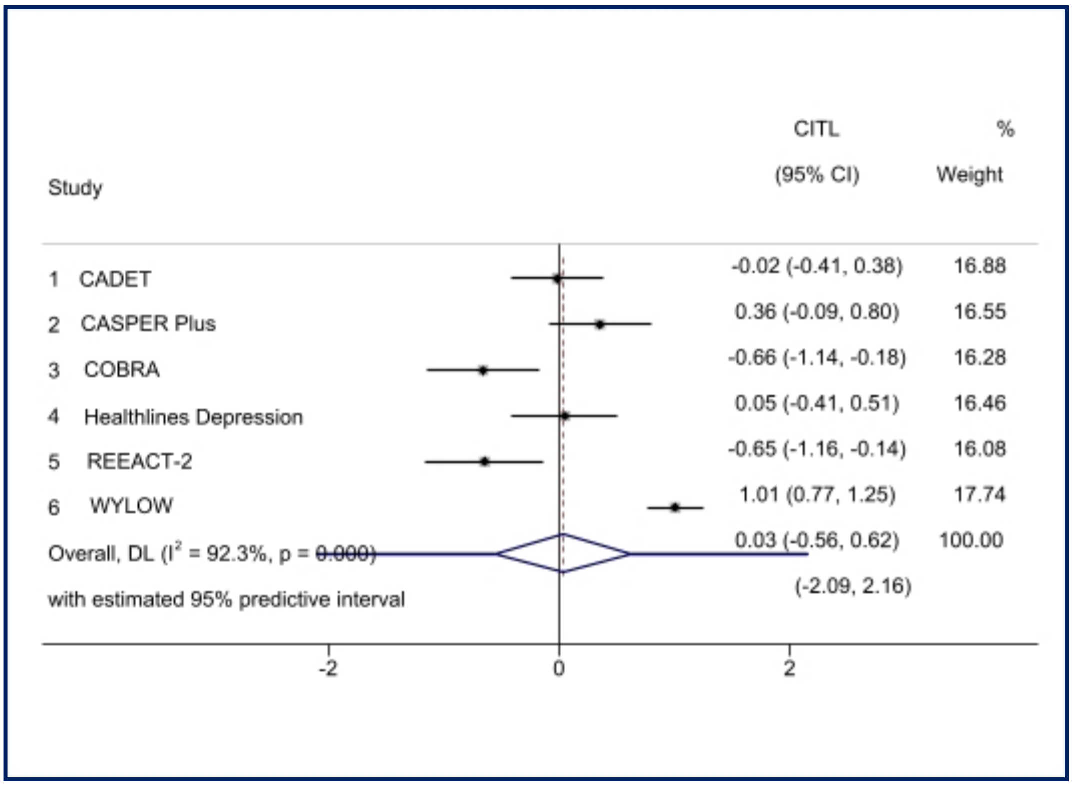

Figure 5.1(c): Pooled CITL for sensitivity analysis (apparent performance)

Table 5.3: Summary of within-cluster and pooled apparent performance statistics for sensitivity analysis

| **Study** | **Number in study** | **C-statistic (95% CI)** | **Calibration slope (95% CI)** | **Calibration-in-the-large (95% CI)** |
|---|---|---|---|---|
| CADET | 158 | 0.59 (0.48 to 0.71) | 0.64 (-0.11 to 1.39) | -0.02 (-0.41 to 0.38) |
| CASPER Plus | 101 | 0.55 (0.42 to 0.68) | 0.26 (-0.59 to 1.11) | 0.36 (-0.09 to 0.80) |
| COBRA | 169 | 0.64 (0.49 to 0.79) | 1.20 (0.19 to 2.22) | -0.66 (-1.14 to -0.18) |
| Healthlines Depression | 110 | 0.64 (0.52 to 0.77) | 1.45 (0.18 to 2.73) | 0.05 (-0.41 to 0.51) |
| REEACT-2 | 159 | 0.66 (0.51 to 0.81) | 1.00 (-0.02 to 2.01) | -0.67 (-1.16 to -0.14) |
| WYLOW | 326 | 0.69 (0.63 to 0.75) | 1.24 (0.79 to 1.69) | 1.01 (0.77 to 1.25) |
| Pooled results | 1023 | 0.65 (0.61 to 0.69) | 0.98 (0.65 to 1.32) | 0.03 (-0.56 to 0.62) |

### 6. Secondary analyses

Following the univariable analysis undertaken as part of the secondary analysis, the analysis here explores relationship status as a predictor in the model. The purpose of this exploratory analysis was to assess the impact of including relationship status as a predictor within the model, given its statistically significant association with relapse on univariable analysis. The same modelling procedures were followed as for the primary analysis. We retained a multilevel logistic regression model with random intercept to preserve the clustering, but did not include the random slope due to convergence issues with a lower sample size. There were 707 participants in the four clusters with data available for the relationship status variable (CADET, COBRA, REEACT and REEACT-2). We initially developed the model in this data, without relationship status (Part 1), to enable me to quantify the effect of adding relationship status (Part 2) and provide a like-for-like comparison.

Part 1: Model development without relationship status

Again, MFPs and mean-centring were applied for continuous predictors (Table 6.1).

Table 6.1: Transformation and mean-centring of continuous predictors following MFP modelling (secondary analysis without relationship status)

| Predictor | Transformation |
|---|---|
| Residual symptoms | X residual_symptoms-4.451202263 |
| Severity | severity-16.53903819 |
| Comorbid anxiety | comorbid_anx_zscore+.1846512682 |

Table 6.2: Multivariable associations (adjusted*) between outcome and predictors for secondary analysis (without relationship status)

| **Predictor** | **Beta coefficient (95% CI)** | **p-value** |
|---|---|---|
| Number of previous episodes | -0.09 (-0.61 to 0.43) | 0.728 |
| Residual symptoms | 0.08 (0.00 to 0.16) | 0.057 |
| Severity | 0.08 (0.02 to 0.14) | 0.008 |
| Comorbid anxiety | -0.14 (-0.39 to 0.12) | 0.287 |
| RCT intervention | -0.37 (-0.81 to 0.07) | 0.102 |

Intercept (baseline risk): -1.60 (95% CI: -2.08 to -1.11)

Standard deviation of random effect on intercept: 0.01

* adjusted for other predictor variables within model

I calculated pooled performance statistics (C-statistic, C-slope and calibration-in-the-large) and also within-cluster statistics to assess heterogeneity in model apparent performance during model development. 95% Prediction intervals were also calculated.

Figure 6.1: Pooled performance statistics (secondary analyses)

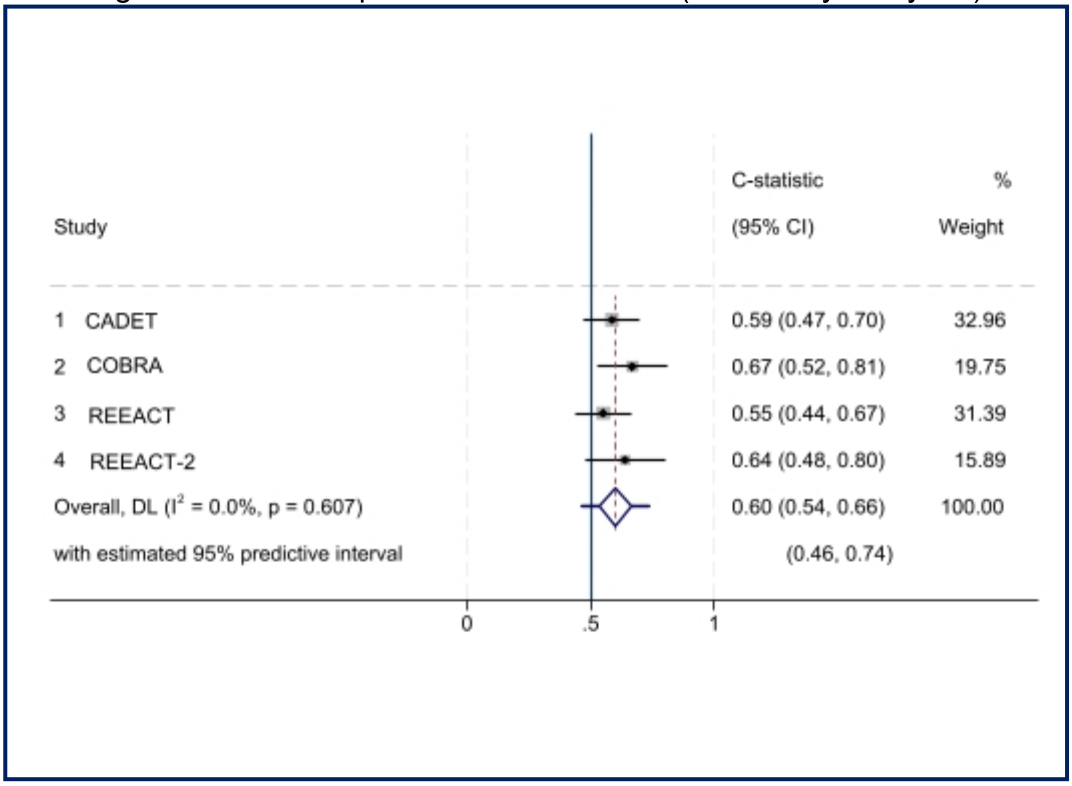

Figure 6.1(a): Pooled C-statistic (apparent performance) for secondary analysis without relationship status

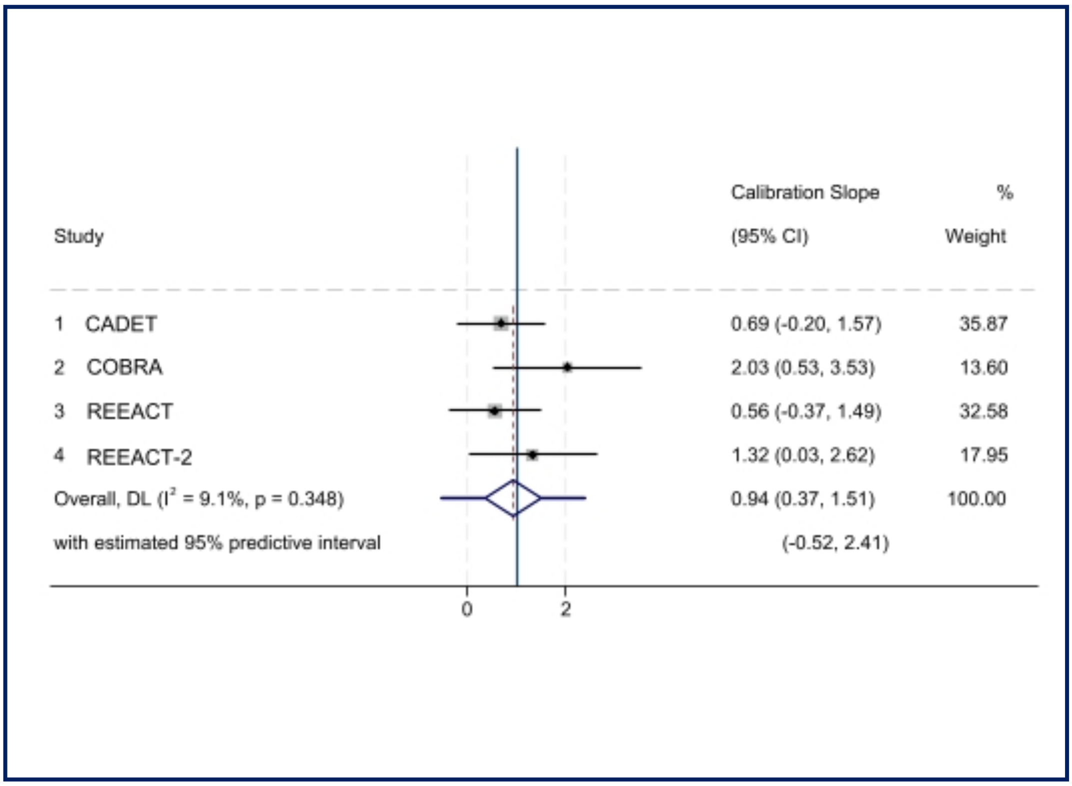

Figure 6.1(b): Pooled calibration slope (apparent performance) for secondary analysis without relationship status

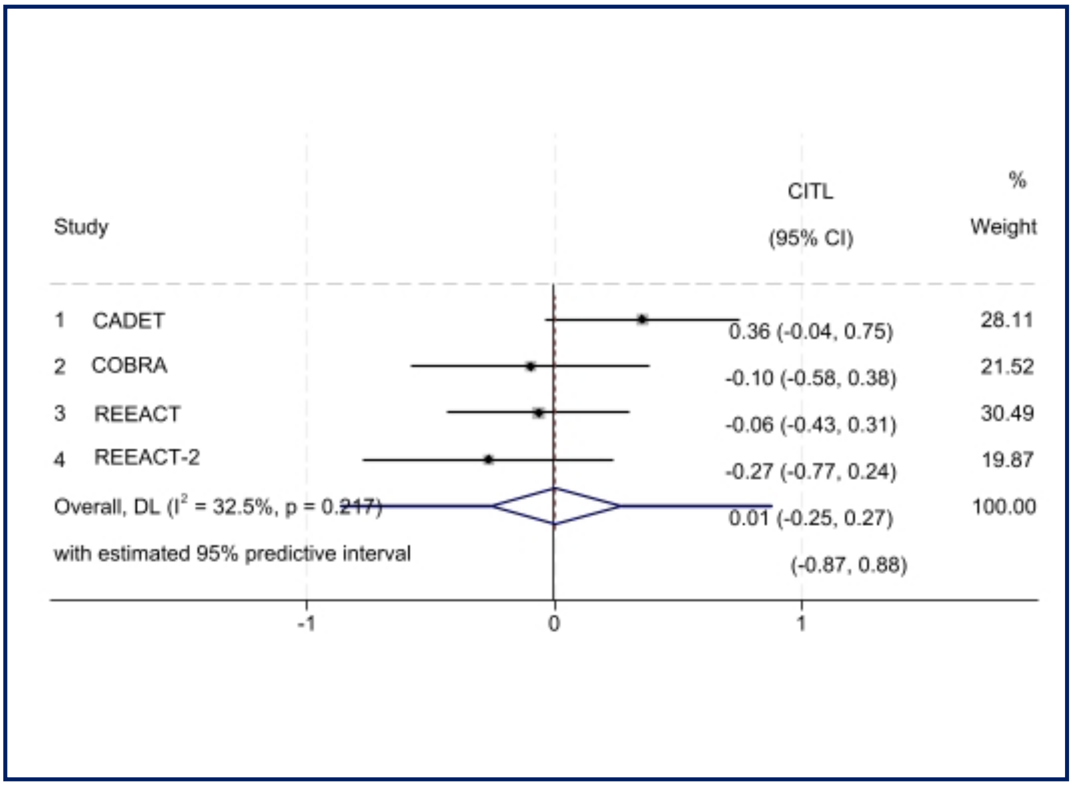

Figure 6.1(c): Pooled CITL (apparent performance) for secondary analysis without relationship status

Table 6.3: Summary of within-cluster and pooled (apparent) performance statistics for secondary analysis without relationship status

| Study | Number in study | C-statistic (95% CI) | Calibration slope (95% CI) | Calibration-in-the-large (95% CI) |
|---|---|---|---|---|
| CADET | 158 | 0.59 (0.47 to 0.70) | 0.69 (-0.20 to 1.57) | 0.36 (-0.04 to 0.75) |
| COBRA | 169 | 0.67 (0.52 to 0.81) | 2.03 (0.53 to 3.53) | -0.10 (-0.58 to 0.38) |
| REEACT | 221 | 0.55 (0.44 to 0.67) | 0.56 (-0.37 to 1.49) | -0.06 (-0.43 to 0.31) |
| REEACT-2 | 159 | 0.64 (0.48 tp 0.80) | 1.32 (-0.03 to 2.62) | -0.27 (-0.77 to 0.24) |
| Pooled results | 707 | 0.60 (0.54 to 0.66) | 0.94 (0.37 to 1.51) | 0.01 (-0.25 to 0.27) |

Part 2: Multilevel logistic regression model with relationship status

Table 6.4: Transformation and mean-centring of continuous predictors following MFP modelling (secondary analysis with relationship status)

| Predictor | Transformation |
|---|---|
| Residual symptoms | residual_symptoms-4.451202263 |
| Severity | severity-16.53903819 |
| Comorbid anxiety | comorbid_anx_zscore+.1855859548 |

Table 6.5: Multivariable associations (adjusted) between outcome and predictors for secondary analysis (with relationship status)

| **Predictor** | **Coefficient (95% CI)** | **P value** |
|---|---|---|
| Number of previous episodes | -0.15<br>(-0.66 to 0.37) | 0.582 |
| Residual symptoms | 0.07<br>(-0.01 0.15) | 0.081 |
| Severity | 0.07<br>(0.01 to 0.13) | 0.020 |
| Comorbid anxiety | -0.12<br>(-0.37 to 0.14) | 0.363 |
| Relationship status | -0.79<br>(-1.23 to -0.34) | 0.001 |
| RCT intervention | -0.40<br>(-0.84 to 0.04) | 0.076 |

Intercept (baseline risk): -1.11 (95% CI: -1.65 to -0.56)

Standard deviation of random effect on intercept: 2.00e—09 (SE = 155.69)

I calculated pooled performance statistics (C-statistic, C-slope and calibration-in-the-large) and also average within-cluster statistics to assess heterogeneity in model apparent performance during model development. 95% Prediction intervals were also calculated.

Figure 6.2. Pooled performance statistics (secondary analyses)

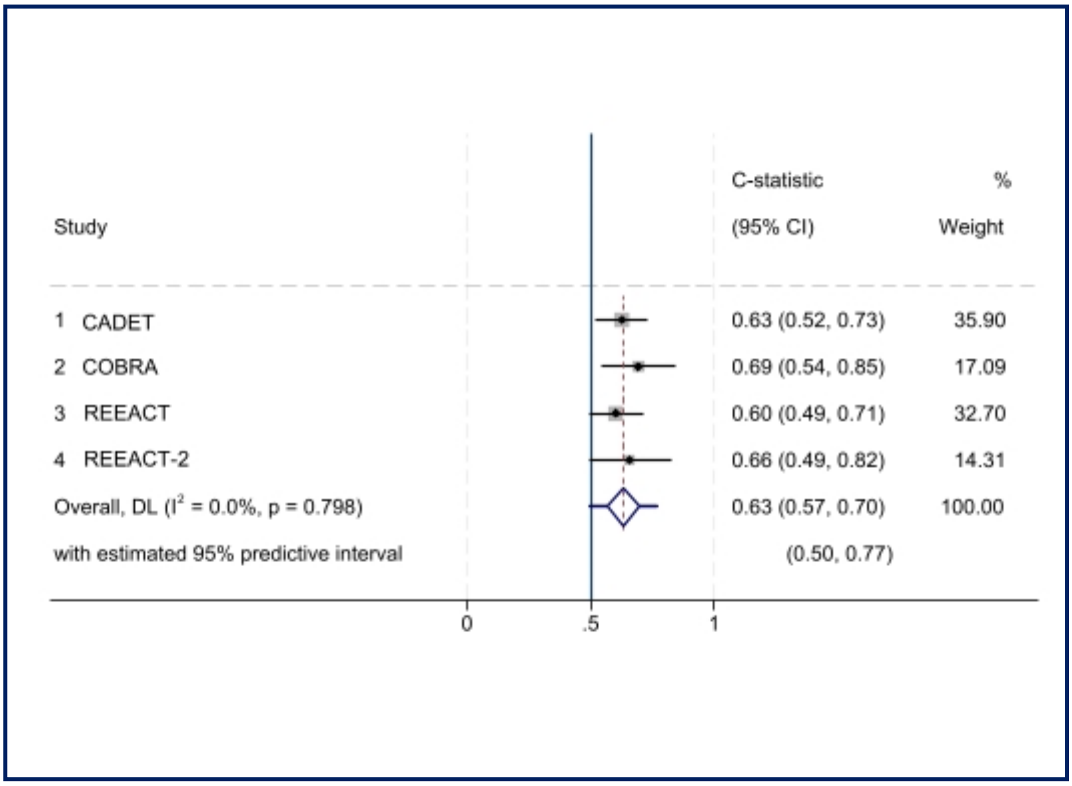

Figure 6.2(a): Pooled C-statistic (apparent performance) for secondary analysis with relationship status

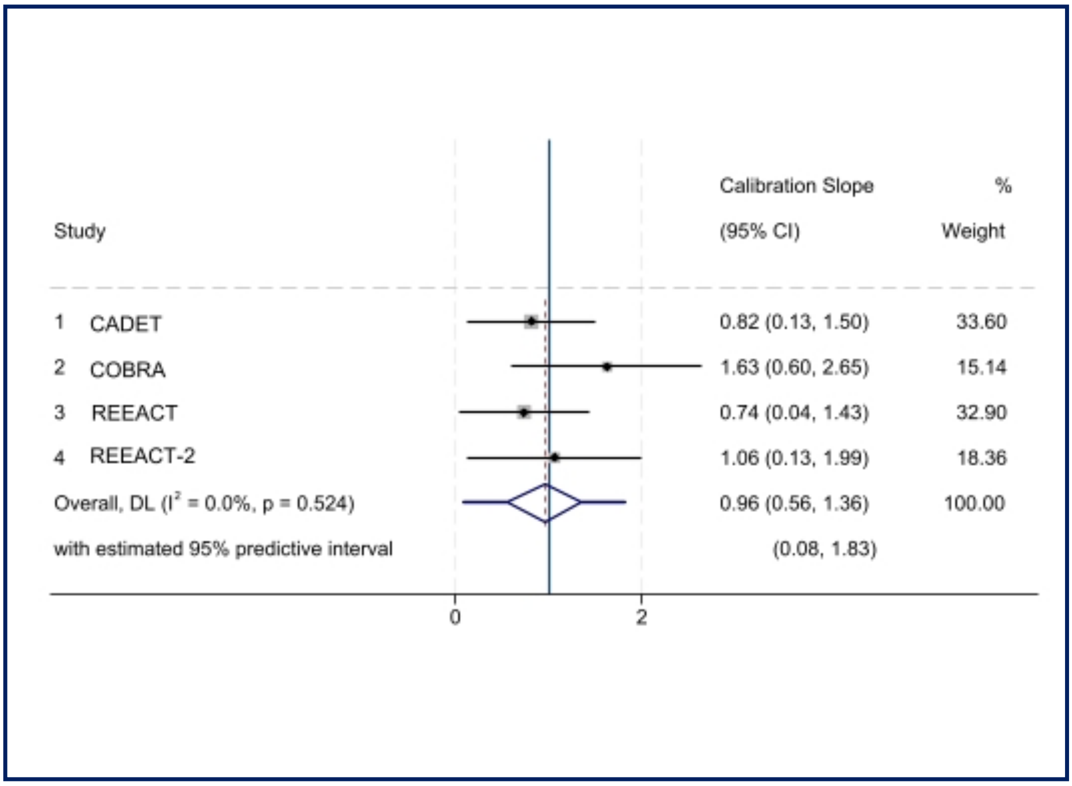

Figure 6.2(b): Pooled calibration slope (apparent performance) for secondary analysis with relationship status

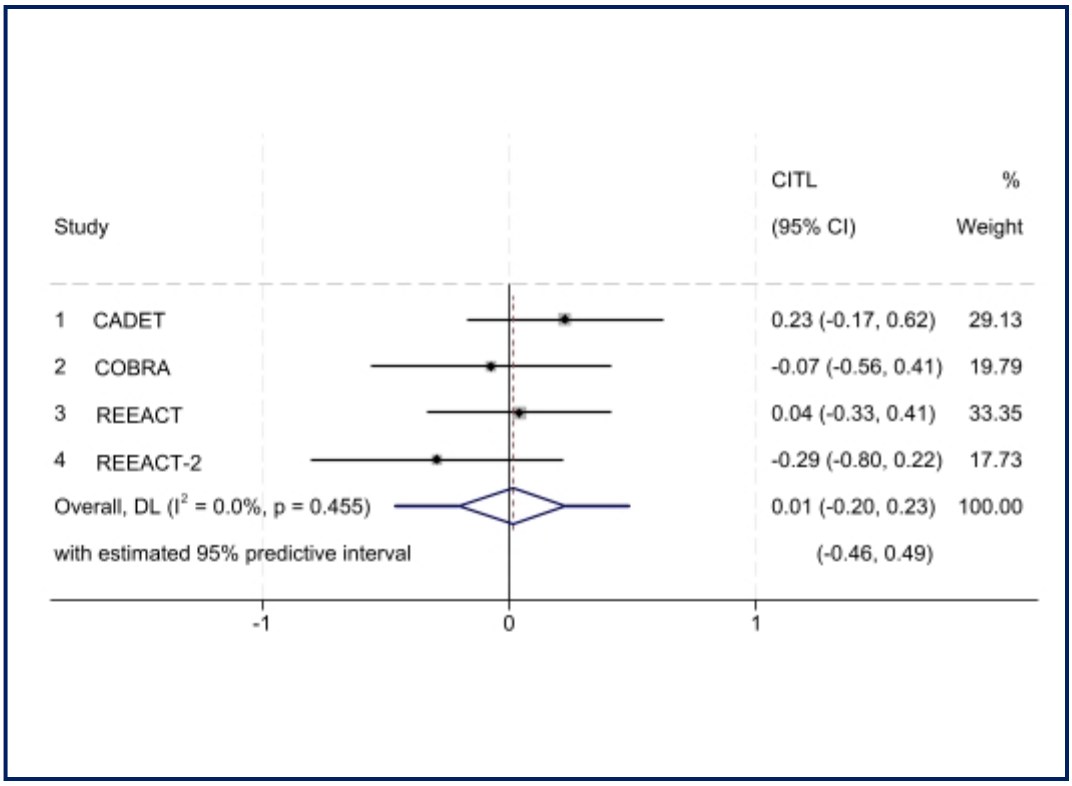

Figure 6.2(c): Pooled CITL (apparent performance) for secondary analysis with relationship status

Table 6.6: Summary of within-cluster and pooled (apparent) performance statistics for secondary analysis with relationship status

| Study | Number in study | C-statistic (95% CI) | Calibration slope (95% CI) | Calibration-in-the-large (95% CI) |
|---|---|---|---|---|
| CADET | 158 | 0.63 (0.52-0.73) | 0.82 (0.13 - 1.51) | 0.23 (-0.17 - 0.63) |
| COBRA | 169 | 0.70 (0.54-0.85) | 1.64 (0.61 – 2.67) | -0.07 (-0.56 to 0.41) |
| REEACT | 221 | 0.60 (0.49 – 0.70) | 0.72 (0.02 - 1.41) | 0.04 (-0.33 to 0.41) |
| REEACT-2 | 159 | 0.66 (0.49 - 0.82) | 1.08 (0.14 - 2.01) | -0.29 (-0.80 to - 0.22) |
| Pooled results | 707 | 0.63 (0.57-0.70) | 0.96 (0.56 - 1.36) | 0.01 (-0.20 to 0.23) |
